# RP-UHPLC/MS/MS Provides Enhanced Lipidomic Profiling of Human Serum in Pancreatic Cancer

**DOI:** 10.1101/2025.09.15.25335762

**Authors:** Zuzana Lásko, Ondřej Peterka, Robert Jirásko, Anna Taylor, Tomáš Hájek, Beatrice Mohelníková-Duchoňová, Martin Loveček, Bohuslav Melichar, Michal Holčapek

## Abstract

**Background:** Pancreatic ductal adenocarcinoma (PDAC) is one of the most lethal cancers, mainly due to the late diagnosis and the lack of reliable biomarkers. Lipidomics provides a promising approach for identifying disease-related alterations, but existing methods are often limited to lipid class profiles with insufficient molecular detail. Reversed-phase ultrahigh-performance liquid chromatography coupled to tandem mass spectrometry (RP-UHPLC/MS/MS) offers the possibility to determine lipids at the fatty acyl/alkyl level. Here, we address the need for a validated quantitative workflow that enables accurate and reproducible lipidomic profiling of human serum in the context of PDAC.

**Results:** We developed and validated an RP-UHPLC/MS/MS method using multiple reaction monitoring, enabling identification of 455 lipid species from 22 subclasses and quantitation of 381 species. The workflow included a response factor correction for sterol esters, which markedly improved their quantification accuracy. The application to serum samples from 54 PDAC patients and 55 healthy controls yielded highly reproducible data, with clear group separation observed in both unsupervised and supervised statistical analyses. Dysregulation was most prominent in sphingolipids and phospholipids. Very long-chain saturated sphingolipids (≥ C22) were downregulated, while some shorter or unsaturated chains showed mild upregulation. Phospholipid alterations were dominated by species containing polyunsaturated fatty acyls, particularly 18:2 and 20:4, with plasmalogens showing the strongest changes. These structurally resolved findings were further supported by gas chromatography – mass spectrometry analysis of fatty acid methyl esters.

**Significance:** This validated workflow provides comprehensive quantitative coverage across 22 lipid subclasses with the structural resolution critical for biological interpretation. The detailed mapping of sphingolipid and phospholipid dysregulation in PDAC demonstrates that only the fatty acyl level annotation reveals molecular signatures that may reflect specific enzymatic activities or pathways. The method delivers a robust platform for biomarker discovery and mechanistic studies in cancer lipidomics.

**HIGHLIGHTS:**

- Validated RP-UHPLC/MS/MS method quantifies 381 lipid species from 22 subclasses.
- Response factors improve the accuracy of sterol ester quantification.
- Lipidomic data enables clear discrimination of cancer and control groups.
- Molecular species resolution reveals hidden lipidomic alterations.
- Sphingolipid dysregulation is mainly determined by *N*-acyl chain composition.

**GRAPHICAL ABSTRACT:** 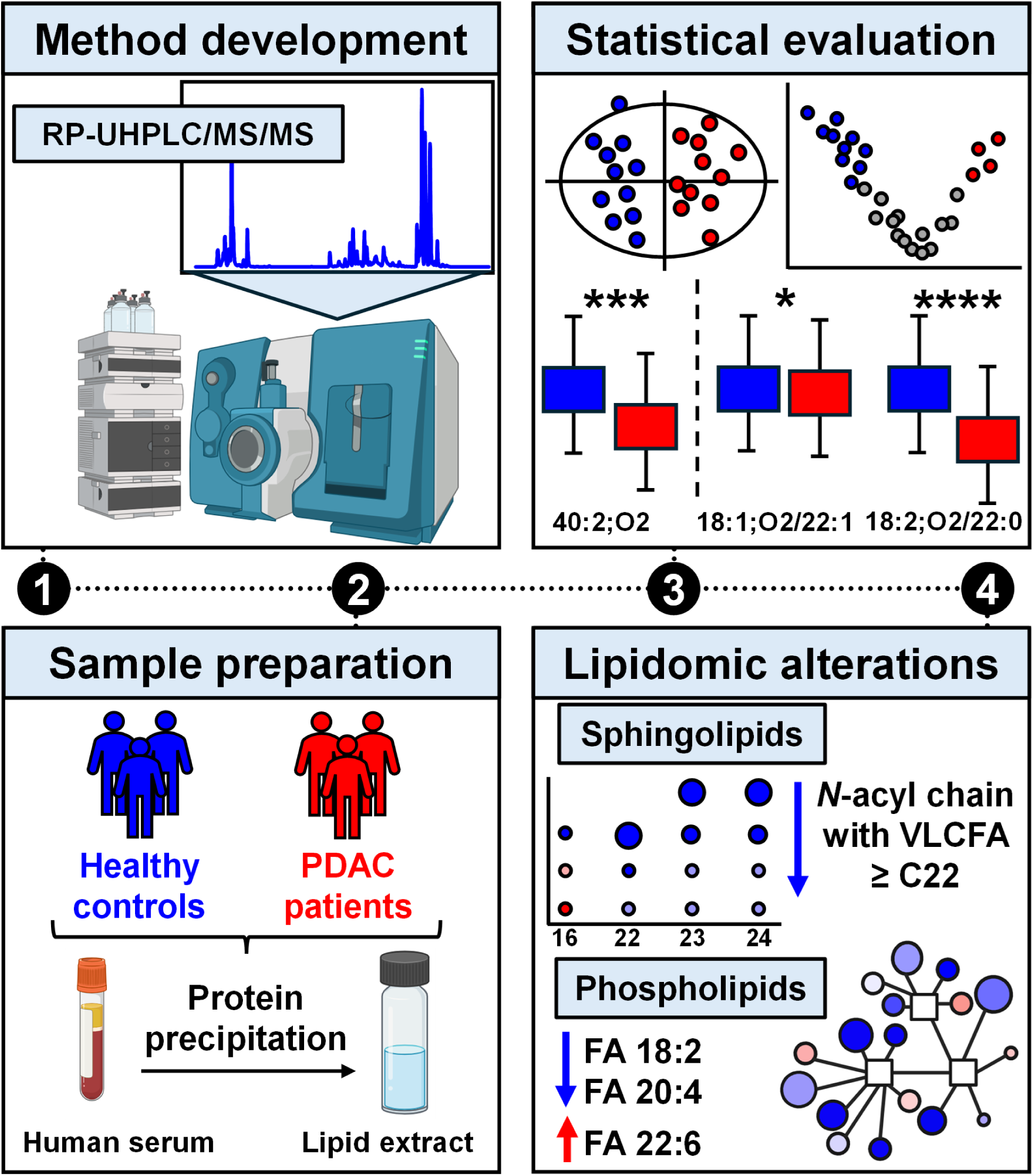

## 1 INTRODUCTION

Pancreatic ductal adenocarcinoma is one of the most aggressive tumors, characterized by high mortality rate. This is primarily due to its frequent diagnosis at an advanced stage, often with metastasis or lymph node involvement, making surgical resection unfeasible. Consequently, PDAC has the lowest 5-year survival rate among all cancers, with an overall rate of 13% and only 3% for distant stage disease [1]. The asymptomatic nature of PDAC in early stages, combined with rapid progression, makes early diagnosis challenging. Current clinical strategies for detecting pancreatic tumors include magnetic resonance imaging, computed tomography, and endoscopic ultrasound, often in combination with measurement of serum concentrations of the Carbohydrate Antigen (CA) 19-9, carcinoembryonic antigen (CEA), or other circulating biomarkers [2–4]. While the integration of imaging techniques with molecular diagnostics has improved diagnostic outcomes, the sensitivity and specificity remain insufficient for early detection. In recent years, significant attention has shifted toward the discovery of novel biomarkers, including exosome analysis [5,6], circulating tumor DNA [7,8], or microRNA [9,10]. Additionally, alterations in lipid profiles have been reported in various tumors, including breast [11–13], lung [14,15], liver [16,17], or kidney [11,18,19] cancers, suggesting that the dysregulation of lipids may play a crucial role in the diagnosis of PDAC [19–21].

Lipidomics focuses on the comprehensive analysis of lipids, including over 49,000 lipid species across 8 lipid categories, in the context of physiological and pathological processes [22]. Changes in lipid profiles are associated with various diseases, including cancer [23], cardiovascular diseases [24,25], and neurodegenerative disorders [26,27]. Examining these changes offers valuable insights into disease mechanisms, with potential for clinical translation. The majority of lipidomic studies utilize mass spectrometry (MS) in combination with various chromatographic techniques based on either lipid class or lipid species separation. Lipid class separation, represented by hydrophilic interaction liquid chromatography (HILIC) [20,28] or ultrahigh-performance supercritical fluid chromatography (UHPSFC) [28–30], allows for high-throughput quantification of endogenous lipids coeluting with internal standards (IS) of the same class. However, this strategy yields only limited information about the molecular structure of individual lipid species. In contrast, reversed-phase ultrahigh-performance liquid chromatography (RP-UHPLC) enables separation of lipids based on their hydrophobicity, fatty acyl chain length, and degree of unsaturation, providing deeper structural resolution and distinguishing of isomeric lipid species [31–33]. Accurate quantification is significantly more complex and often requires the use of multiple IS or concentration correction using response factors. Consequently, relative quantification using one or more IS are generally preferred, as this approach is often adequate for comparative analyses and for detecting relative changes in lipid concentrations across different samples.

Human serum and plasma are the most widely used biological matrices in biomarker discovery due to their accessibility and ability to reflect systemic physiological changes. However, the wide concentration range of lipid species in these matrices requires sensitive and selective analytical techniques. Multiple reaction monitoring (MRM) enables targeted detection and quantification of lipids with high specificity and reproducibility, even at trace levels [33–35].

In this study, we present the development and application of the targeted RP-UHPLC method coupled with low-resolution MS operating in MRM mode for the quantitative lipidomic analysis of human serum samples. Compared to our previously used lipid class methods, this approach provides significantly improved structural resolution of lipid species, allowing for more accurate characterization of lipidomic alterations associated with PDAC. Comparison of lipid profiles of serum samples from healthy individuals and PDAC patients, we aim to identify structurally resolved lipid markers with the potential diagnostic relevance.

## 2 MATERIAL AND METHODS

### 2.1 Chemicals and standards

Methanol, butanol, 2-propanol, acetonitrile, and formic acid (all in LC/MS grade) were purchased from Honeywell (Charlotte, North Carolina, USA). LiChrosolv chloroform (stabilized with 2-methyl-2-butene) and ammonium formate (for MS, ≥99.0%) were obtained from Merck (Darmstadt, Germany). Deionized water was produced using a Milli-Q Reference Water Purification System (Molsheim, France). IS were obtained from Merck (Darmstadt, Germany), Nu-Chek (Elysian, MN, USA), and Avanti Polar Lipids (Alabaster, AL, USA). Stock solutions of all lipid standards were prepared in methanol/chloroform (1:1, *v/v*). Final concentrations of all IS in the mixture (IS-mix) are provided in Table S1. Cholesteryl ester standards used for the calculation of response factors were obtained from Nu-Chek, with concentrations in the stock solution listed in Table S2.

### 2.2 Biological samples

Human serum samples were obtained from 54 PDAC patients (aged 36–80 years; body mass index (BMI) 17–41) and 55 healthy volunteers (aged 25–74 years; BMI 20–32), provided by Palacký University Hospital Olomouc, Czech Republic. The clinical information for all subjects is summarized in Table S3. Quality control (QC) samples were prepared by pooling equal aliquots from both healthy controls and PDAC groups and were injected at regular intervals (every 20 injections) to monitor the instrumental stability throughout the analytical sequence. All samples were stored at −80 °C prior to the lipidomic extraction. The study was approved by the institutional ethics committees in accordance with the Declaration of Helsinki, and written informed consent was obtained from all participants. The NIST SRM 1950 human plasma was used as a reference material for the quantitative analysis and the comparison with published lipid concentrations.

### 2.3 Sample preparation

The lipidomic extraction from human serum was performed using a protein precipitation method [33]. In this protocol, 25 µL of serum was mixed with 20 µL of IS-mix (Table S1) and deproteinized using 250 μL of butanol/methanol solution (1:1, *v/v*). The mixture was sonicated in an ultrasonic bath at 25 °C for 15 min and subsequently centrifuged (Hettich EBA 20) at 6,000 rpm (3468×g) for 15 min at room temperature. The supernatant was purified using a plastic syringe fitted with a 0.25 µm cellulose membrane filter, which was then rinsed with 500 µL of the butanol/methanol solution (1:1, *v/v*). The combined filtrate was evaporated to dryness under a gentle stream of nitrogen at 35 °C. Prior to the analysis, the residue was reconstituted in 250 µL of chloroform/methanol mixture (1:1, *v/v*) and vortexed for 1 minute to ensure the complete dissolution.

### 2.4 Fatty acyl methyl esters preparation

For the transesterification of lipids, 25 µL of human serum (30 samples from healthy volunteers and 30 samples from PDAC patients) spiked with 10 µL of the IS FA 19:1 (10Z) (0.3 mg/mL in chloroform/methanol, 1:1, *v/v*) were processed by protein precipitation (Chapter 2.3). The precipitate was reconstituted in 250 µL of chloroform/methanol (1:1, *v/v*). Then, 100 mg of this extract was mixed with 1.5 mL of sodium methoxide solution in methanol (0.25 M), vortexed for 1 min, and incubated in a heating block at 60 °C for 20 min. During the esterification reaction, the vial was shaken briefly every five minutes. After cooling to ambient temperature, 1 mL of saturated sodium chloride solution and 2 mL of hexane were added, and the vial was vortexed for 1 min. The organic phase containing fatty acyl methyl esters (FAMEs) was collected, evaporated under a gentle stream of nitrogen, and reconstituted in 100 µL of hexane for gas chromatography–mass spectrometry (GC/MS) analysis.

### 2.5 RP-UHPLC/MS/MS conditions

Sample analyses were performed on an Agilent 1290 Infinity series liquid chromatograph (Agilent Technologies, Waldbronn, Germany) coupled to low-resolution hybrid quadrupole-linear ion trap (QTRAP) 6500 mass spectrometer (SCIEX, Framingham, MA, USA). The chromatographic separation was performed on Acquity UPLC BEH C18 column (150 × 2.1 mm, 1.7 μm, Waters) equipped with Acquity UPLC BEH C18 Van Guard Pre-column (50 × 2.1 mm, 1.7 μm, Waters). The column temperature was set to 55 °C. The mobile phase consisted of solvent A: acetonitrile/water (60:40, *v/v*) and solvent B: acetonitrile/2-propanol (10:90, *v/v*), both containing 0.1% formic acid and 5 mM ammonium formate. The gradient elution program was applied as follows: 0 min – 35% B, 8 min – 50% B, 21-23 min – 95% B, and 24-25 min – 35% B, followed by 1 min post-run equilibration. The flow rate was set to 0.35 mL/min, and the sample volume of 1 µL was injected. To minimize carry-over and ensure consistent system performance, the injector needle was washed after each injection using the mixture of isopropanol/methanol/chloroform (4:2:1, *v/v/v*) containing 5% water.

The mass spectrometric analysis was performed in the positive ion mode using a Turbo V electrospray ionization source. The capillary voltage was set to 5.5 kV, and the source temperature was maintained at 250 °C. The curtain gas was set to 20 psi, nebulizer gas to 50 psi, and turbo gas to 50 psi. The entrance potential was set to 10 V, and the collision cell exit potential to 15 V. Specific MRM transitions were optimized for each analyte using IS-mix, including adjustments to retention time windows, dwell weights, collision energy, and declustering potential. Optimized MRM parameters for the individual lipid subclasses and IS are summarized in Table S4. Data acquisition and instrument control were performed using Analyst software (version 1.6.2; Sciex).

### 2.6 GC/MS conditions

FAMEs were analyzed using an Agilent 7890A gas chromatograph coupled to an Agilent 5977A mass spectrometer with electron ionization (both Agilent Technologies, Waldbronn, Germany) using a TRACETM TR-FAME column (60 m × 0.25 mm, 0.25 µm, Thermo Scientific, Waltham, USA). GC conditions were set as follows: injection volume 5 µL, injector temperature 250 °C, split ratio 1:1, helium flow rate 1.5 mL/min. The temperature program started at 80 °C, increased to 150 °C at 10 °C/min, then to 235 °C at 3 °C/min, and was held at 235 °C for 20 min. The ion source and quadrupole temperatures were set to 250 °C and 150 °C, respectively. The mass spectrometer operated in the scan range *m/z* 50–600 with an acquisition rate of 5 scans/s and a solvent delay of 7 min.

### 2.7 Data processing

Raw data files acquired from the RP-UHPLC/MS system were exported in the .wiff format and processed using the Skyline software (MacCoss Lab Software, University of Washington, Seattle, WA, USA) [36]. Lipid concentrations were calculated based on the ratio of the peak height of each lipid species to the peak height of a corresponding IS, multiplied by the known concentration of that standard. Only lipid species quantified in at least 80% of all samples were included in the dataset for further analysis. For missing values, zero filling was applied by setting 80% of the minimum measured concentration of the respective lipid species across all samples. In addition, all values below the defined limit of quantitation (LOQ) were removed from the dataset. Lipid species with a relative standard deviation (RSD) exceeding 20% in repeated analyses of QC samples and NIST SRM 1950 plasma were also excluded. Data from GC/MS measurements were processed and interpreted using Agilent MassHunter software (Agilent Technologies, USA).

### 2.8 Statistical analysis

Statistically significant differences in lipidomic profiles between healthy controls and cancer patients were evaluated using univariate methods, including a two-sided Welch’s t-test (assuming unequal variances) and fold change (FC) analysis. Lipid species with the FC (tumor/normal) greater than 1.2 or less than 0.8 (i.e., a relative change exceeding 20%) and a *p*-value < 0.05 were considered statistically significant. Multivariate statistical analysis was performed in the R software environment (version 4.4.1) and included principal component analysis (PCA), orthogonal partial least squares discriminant analysis (OPLS-DA), and visualization methods, such as box plots and volcano plots. Prior to the analysis, lipid concentrations were log-transformed and Pareto-scaled to enhance the data normalization. The variable importance in projection (VIP) scores were calculated using SIMCA software (version 13.0.3; Umetrics, Sweden). Lipid species with VIP values greater than 1 were considered statistically relevant contributors to the group separation. Heatmap combined with clustering analysis were used to visualize lipid concentrations in biological samples, with a color scale highlighting differences across groups. The S-plot generated from OPLS-DA model was applied to identify the most dysregulated lipid species, while box plots were used to display concentration changes of selected lipids. The statistical significance was further confirmed using the non-parametric Mann–Whitney U test, with significance levels denoted as follows: ns (non-significant; *p* > 0.05), * (*p*=0.05–0.01), ** (*p*=0.01–0.001), *** (*p*=0.001–0.0001), and **** (*p* ≤ 0.0001). The Cytoscape software (version 3.8.2) was used for the construction of network maps.

## 3 Results and discussion

### 3.1 Method development

RP-UHPLC/MS is among the most widely used approach in lipidomics due to its ability to separate and characterize a broad spectrum of lipid species, including isomeric forms that differ in fatty acyl chain length or degree of unsaturation. In our previous untargeted method [32], RP-UHPLC coupled with high-resolution MS was primarily used for highly confident lipid identification based on mass accuracy within 5 ppm tolerance, characteristic fragment ions in MS/MS, and retention dependences. However, the present work focuses on the quantitative lipidomic analysis and profiling in clinical cohorts, where low-resolution instruments operated in MRM mode offer higher sensitivity, selectivity, and reproducibility.

For the method optimization, we used the IS-mix (Table S1), with most lipid subclasses represented by two standards. The chromatographic settings in this study were taken from our previously published RP-UHPLC/MS/MS method [32] optimized for high resolution MS detection, but for the transition to the low-resolution mass spectrometer, it was necessary to adjust the mass spectrometric parameters. This included the selection of the most intense precursor and fragment ions for each lipid subclass, and the optimization of collision energies and declustering potentials. The source temperature, capillary voltage, and gas pressure settings were set based on our previously published lipidomics methods [12,19]. Precursor and characteristic fragment ions for each lipid subclass were systematically selected using full-scan and MS/MS experiments, while collision energies and declustering potentials were optimized in MRM mode with ramps from 5 to 80 V and 0 to 300 V. Values giving the highest signal responses were chosen as optimal, except for phosphatidylcholines (PC) and cholesterol esters (CE), where slightly lower collision energies were applied to prevent detector saturation caused by their high concentrations. Dwell weights were set for individual lipid subclasses according to the concentrations of endogenous lipid species with priority given to less abundant classes, such as phosphatidylinositoles (PI), lysophosphatidylinositoles (LPI), and sulfatides (SHexCer). All final MS parameters, MRM transitions, and retention times for IS are summarized in Table S4.

### 3.2 Lipid species separation and their identification

A pooled human serum sample was used for lipid identification and the qualitative analysis was performed using a home-prepared lipid database in combination with characteristic neutral loss (NLS) and precursor ion scanning (PIS) modes. However, SHexCer were detected primarily using MRM transitions due to their low ion intensities in positive ion mode. The identification was supported by evaluation of retention behavior, including the construction of polynomial dependencies of retention times on fatty acyl chain length and degree of unsaturation (data not shown). Moreover, the identification is in the line with our previous untargeted method [32], but new methods bring higher sensitivity and more identified lipid species. The typical chromatographic behavior of individual lipid subclasses is shown in the total ion chromatogram of human serum sample (Figure 1A) and in extracted ion chromatograms of individual IS (Figure S1). Lipid nomenclature and shorthand notation followed the recommendations of Liebisch *et al.* [37], where species level reflects the total number of carbon atoms and double bond(s), while the fatty acyl/alkyl level provides detailed information about the composition of individual fatty acyls. The *sn*-positions of phospholipid species were annotated based on our previous work [32], and only the prevailing isomer (determined by MS/MS experiments) is reported.

**Figure 1.**
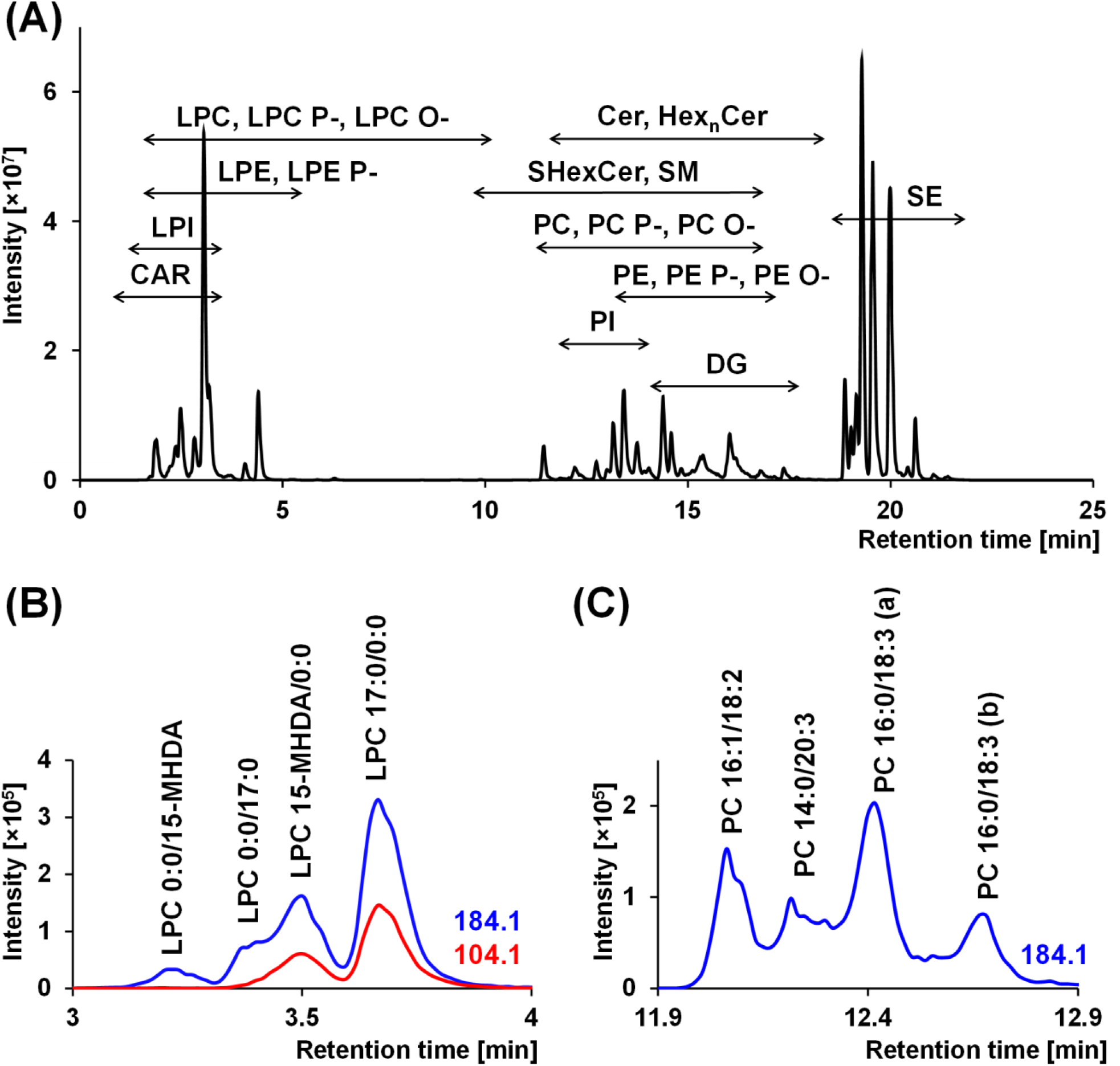
Chromatographic separation of: (A) human serum sample, (B) LPC 15-MHDA and LPC 17:0 detected using PIS 184 and PIS 104, and (C) PC 34:3 isomers measured by RP-UHPLC/MS/MS in the positive ion mode. Peak identification is based on data from our previous article [32], where annotation was determined according to the prevailing isomer.

The most monitored adduct ions were [M+H]^+^ or [M+NH_4_]^+^ and class-specific fragmentations were applied for the identification. All sphingolipids, represented by sphingomyelins (SM), ceramides (Cer), n-hexosylceramides (HexnCer), and SHexCer, were monitored using PIS of the sphingoid base. For SM, although the most intense fragment is *m/z* 184, the less intense sphingoid base transitions were used to obtain more detailed structural information on the molecular composition. Lysophosphatidylcholines (LPC) species were detected using PIS 184 and 104, with *m/z* 104 being characteristic for fatty acids bound at the *sn-1* position (Figure 1B) and for plasmalogen-/ether-linked LPC P-/LPC O-species. Notably, LPC P- and LPC O-species exhibited significantly higher intensity for the *m/z* 104 fragment. Phosphatidylcholines (PC, PC P-, PC O-) were measured using PIS 184. Lysophosphatidylethanolamines (LPE and LPE P-) and phosphatidylethanolamines (PE and PE O-) were identified by NLS 141, while plasmalogen PE P-species were detected using acyl-specific PIS. LPI and PI were analyzed by NLS 277. Although these transitions for phospholipids are not specific for individual isomers (Figure 1C), the fatty acyl composition was assigned based on the previous experimental experience and literature data [31–33,38]. Diacylglycerols (DG) were identified using NLS of the corresponding fatty acids and sterol esters (SE) were monitored using PIS 369, 367, and 397. The *m/z* 369 fragment is common to both cholesterol and lathosterol esters, with cholesterol esters presumed to be the main source due to their high abundance in human plasma. The *m/z* 367 transition is characteristic of desmosterol and 7-dehydrocholesterol esters, while *m/z* 397 corresponds to sitosterol esters. Carnitines (CAR) were analyzed using the characteristic PIS 85. All characteristic NLS and PIS transitions for the analyzed lipid subclasses are summarized in Table S4, while Table S5 provides the corresponding data for all lipid species detected in pooled human serum.

In total, 455 lipid species from 22 lipid subclasses were identified in pooled human serum and the number of identified lipid species was compared with the previously published data, as summarized in Table S6. This comparison included methods employing RP separation [32,33], as well as comprehensive inter-laboratory studies that utilized a range of analytical approaches [39–42]. Compared to our previously published qualitative method using high-resolution MS [32], the current approach represents an increase of 83 species. The most notable improvements were observed for SHexCer (17 species, previously not detected) and SE (47 *vs.* 13), due to the higher sensitivity of the MRM approach. This also enabled the detection of sterol esters with 27:2 and 29:1 sterol backbone, which has not been reported previously. The substantial increase was also achieved for PE P-species (41 *vs*. 27), enabled by acyl-specific transitions. In contrast, lower number of Hex3Cer species were detected (1 *vs*. 7) due to their high molecular masses exceeding the calibration range of the instrument. Triacylglycerols (TG) were not analyzed in this study due to the complexity of their structure and the challenges of accurate quantification on low-resolution instruments. Although TG quantitation is most commonly based on the neutral loss of one fatty acyl chain, this approach carries a high risk of misquantification, as better chromatographic separation is required to resolve individual TG isomers. In addition, the composition of TG is strongly influenced by lifestyle factors, particularly diet, and their lipidomic profiles change considerably over time. For these reasons, TG have limited potential as cancer biomarkers. Monoacylglycerols and sterols, identified on high-resolution instrument, were not quantified due to the absence of suitable MRM transitions. For these lipid classes, derivatization strategies may represent a useful option to improve sensitivity and enable quantification using LR-MS platforms [34].

In the inter-laboratory comparison, our method provides the lipidomic coverage largely consistent with most previously reported approaches, exceeding that of Bowden *et al*. [42] (247 species), Ghorasaini *et al*. [40] (207 species), Mandal *et al*. [39] (318 species), and was broadly comparable with Quehenberger *et al*. [41] (382 species) and Huynh *et al.* [33] (559 species). Notably, higher numbers were obtained for sterol esters (47 species *vs*. 19–33 in other approaches) and for SHexCer (17 *vs*. 6), highlighting the sensitivity of the applied MRM-based workflow. In contrast, the number of plasmalogens and ether-linked PC/PE species was lower compared to Huynh *et al.* [33] (137 *vs*. 70). Differences in the number of lipid species detected within individual classes across studies may in part be attributed to variations in LC/MS platforms, extraction procedures, and methodological focus. Overall, lipid the coverage achieved by our workflow is largely consistent with that of other published methods.

### 3.3 Method validation

The method validation is crucial to ensure accurate and reliable results due to a large number of various lipid classes and subclasses with a wide range of concentrations. For this purpose, pooled human serum samples (25 µL) were spiked with IS-mix either before or after the extraction process. Each IS was carefully selected to represent lipid subclasses, based on the assumption that it exhibits similar behavior to endogenous lipids in human serum. The validation was performed in accordance with standard bioanalytical guidelines [28] and included the evaluation of calibration range, limit of detection (LOD), LOQ, precision (repeatability and instrument precision), accuracy, extraction recovery, selectivity, carry-over, and matrix effect. Calibration curves (Figure S2) were constructed based on investigation of fourteen concentration levels, with each level extracted in triplicate. The linearity was evaluated based on the coefficient of determination (R^2^), which exceeded 0.998 for all analyzed IS. The LOD was defined as the lowest IS that could be repeatedly and reliably detected. The LOQ was defined as the lowest concentration of IS that could be quantified with a maximum relative standard deviation (RSD) of 20%. The results of calibration ranges, LOD, and LOQ for all IS are summarized in Table S7A.

The carry-over was evaluated by injecting a blank solvent sample immediately after the highest concentration level of the calibration curve. The residual signal in the blank was required to remain below 20% of the signal at LOQ. To minimize analyte transfer between injections, the system was programmed to perform thorough washing of the injector needle and needle seat during each run (initiated at 23 min of the gradient). Precision, accuracy, extraction recovery, and matrix effects were evaluated at three concentration levels selected based on the calibration curves: low (LL; 10 µL spike), medium (ML; 20 µL spike), and high (HL; 40 µL spike). Repeatability (intra-day precision) was evaluated using six independently prepared pooled serum extracts, while instrumental precision was determined by analyzing six consecutive injections of pooled serum samples. For both parameters, RSD remained below 15% across all tested concentration levels. Accuracy was assessed by calculating concentrations from the calibration curves compared to the theoretical spiked values. Most IS exhibited RSD within 15%; exceptions were observed for LPE 14:0 (17%) and Gb3 18:1;O2/17:0 (16%) at the low concentration level. Overall, the analytical method demonstrated good accuracy and precision, particularly at the medium concentration level, which is commonly used for lipid quantification in biological samples. For the evaluation of matrix effect, extraction recovery, and selectivity, individual serum samples from six randomly selected clinical cohort participants were analyzed. Extraction recovery was evaluated by comparison of the signal intensities of samples spiked with the IS-mix before extraction to those spiked after extraction. All IS showed relatively high extraction yields, particularly at the ML (78–103%). Matrix effect was determined by calculating the ratio of signal intensities from individual serum samples spiked with the IS-mix after extraction to those obtained from equivalent concentrations of neat IS solutions. The matrix factor (MF) was calculated for each lipid subclass as the ratio of peak area in matrix to peak area in solvent. Most MF at all tested concentration level were <1, indicating reduced ionization efficiency. However, the RSD values for all standards remained within 15%, which means that the matrix effect is not considered as an exclusion criterion. Selectivity was evaluated by comparing the responses of each sample with and without the addition of the IS-mix at a LL after extraction. The intensity of any endogenous interference in the non-spiked matrix was required not to exceed 15% of the signal intensity of the corresponding IS. Almost all IS fulfilled this criterion, except for LPE 14:0, CAR 14:0 d9, CAR 20:4 d4, and SHexCer 18:1;O2/12:0, which show interference values of 15%, 25%, 31%, and 34%, respectively, and were therefore excluded from further quantitative evaluation. Notably, the results also indicate that deuterated IS exhibit higher selectivity compared to exogenous analogues, which is evident from the obtained data. The results of all these parameters are summarized in Table S7B.

### 3.4 Quantitation of lipid species in NIST SRM 1950 plasma

The validated RP-UHPLC/MS/MS method was applied to the quantitative analysis of lipids in NIST SRM 1950 human plasma spiked with IS-mix before the extraction to verify accuracy of the determined concentrations with literature values [33,39–42], resulting in the quantification of 381 lipid species from 21 lipid subclasses (Table S8). For most lipid subclass, two IS were employed (expect of LPI, PE P-, PC P-, SHexCer, and SE), and concentrations were calculated using both peak areas and peak heights. The correlation plot (Figure 2A) demonstrates that both quantification approaches provide comparable results. For lipids, particularly phospholipids (PC, PE, and PI), where isomeric species were not resolved on the baseline and peak area integration is therefore more challenging, the calculation based on peak height brings more reliable and reproducible quantitation. For LPC, both isomers were quantified (when chromatographically separated) using the common PIS 184 and the PIS 104 transition specific for the *sn-1* form. Figure 2B shows that both transitions yielded comparable results, while the PIS 104 additionally enabled quantification of the *sn-1* isomer in cases where the *sn-1* and *sn-2* forms were not chromatographically resolved. However, a difference in concentration was observed for LPC P- and LPC O-species, where the comparison with literature data indicated that the PIS 104 provided more accurate results. This transition also exhibits substantially higher intensity for these subclasses. DG were quantified based on the NLS of both fatty acyl chains and the intensities obtained for the individual fatty acyl losses were summed. The resulting values were then used for the calculation of lipid concentrations.

**Figure 2.**
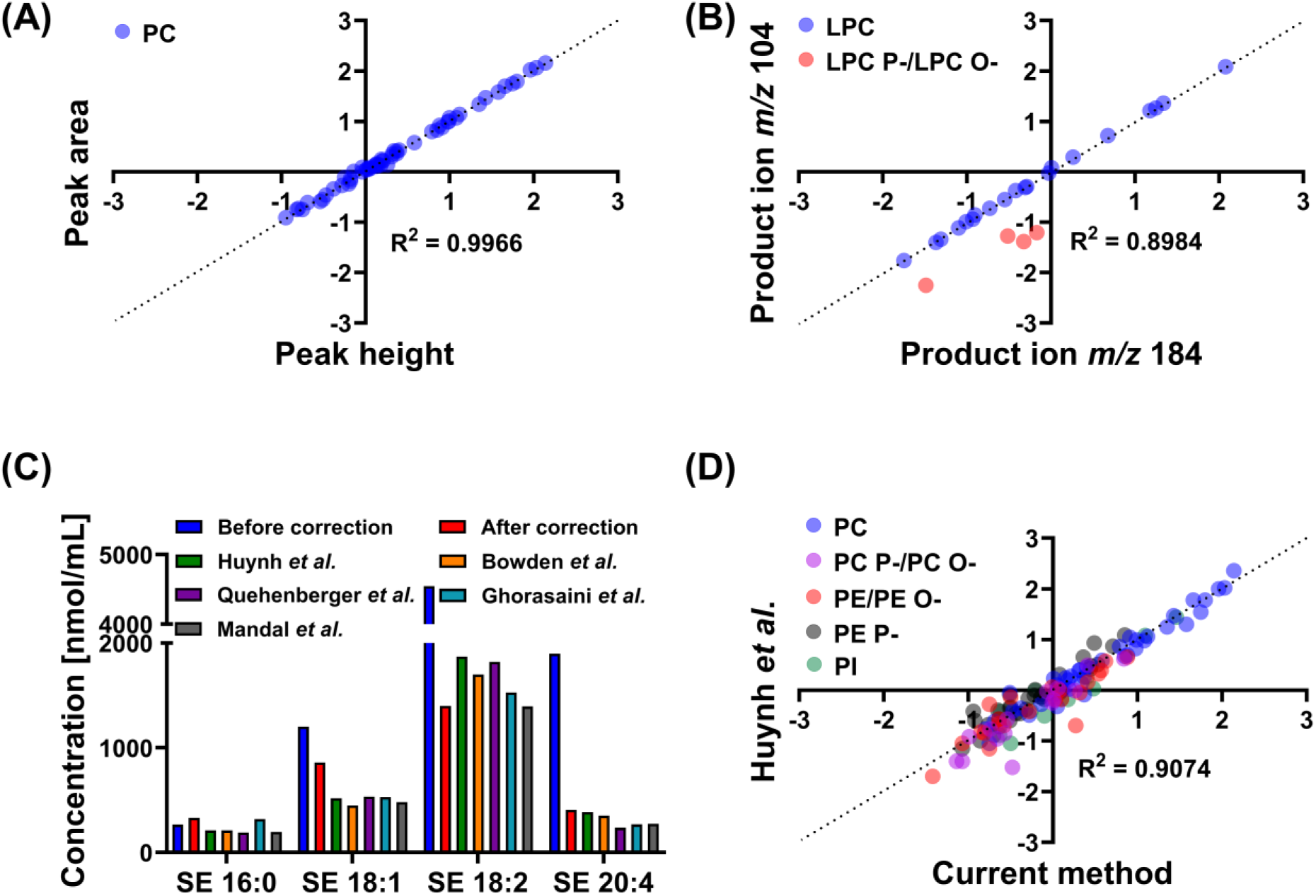
Comparison of final lipid concentrations in NIST SRM 1950 human plasma. (A) Correlation plots of PC concentrations calculated using peak height and peak area. (B) Correlation plots of LPC concentrations obtained using PIS 184 and PIS 104. (C) Comparison of SE concentrations corrected using response factors and literature values. (D) Correlation plots of phospholipids concentrations measured by current method and by Huynh *et al.* [33] using RP-UHPLC/MS/MS.

The accurate quantification of SE is challenging, as these lipids exhibit differences in ionization efficiencies in MS depending on their structure (i.e., the number of double bonds and the fatty acyl chain length. Therefore, the use of specific response factors (RF) is essential to achieve their accurate quantification [43,44]. In this study, RF values were experimentally determined for 14 cholesteryl ester standards. The values were calculated as the ratio of the calibration curve slope of each standard relative to the reference compound CE 19:0 (RF = 1), according to the equation: RF(CE X:Y) = a(CE 19:0) / a(CE X:Y), where *a* presents the slope of the calibration curve. An additional correction was applied to improve accuracy by accounting for the distinct ionization behavior of the deuterated IS CE 16:0-d7. The experimentally obtained RF values (Table S2) showed systematic trends depending on fatty acyl chain length and unsaturation (Figure S3). Response factors for other cholesteryl esters not included in the available standards were extrapolated from the regression equation. The significant effect of this correction is demonstrated in Figure 2C, which compares uncorrected data, RF-corrected data, and literature values. These results clearly highlight that the application of response factors is indispensable for the reliable and accurate quantification of the SE lipid class.

Quantitative results obtained for NIST SRM 1950 plasma were compared with datasets reported in previous studies employing RP-UHPLC/MS/MS [33], and with results from studies employing various analytical techniques [39–42]. Overall, our concentrations correlate with the literature values, showing only minor deviations that can be attributed to differences in extraction protocols, IS compositions, analytical approaches, instrumentation, and data processing workflows between studies [45]. The largest differences were observed in comparison with Quehenberger *et al*., [41], particularly for LPE, LPC, PE, and PC classes. However, the dataset of Quehenberger *et al.* also shows considerable differences when compared with other inter-laboratory studies, which may partly explain these deviations. In addition, some of these methods do not report concentrations at the fatty alkyl/acyl level, but only as summed values and, therefore, the most appropriate comparison is with the study by Huynh *et al.* [33] employing also an RP-UHPLC/MS/MS approach in combination with a low-resolution instrument. Figure 2D illustrates the correlation (R^2^ = 0.9074) between all phospholipid concentrations obtained by current method and those reported by Huynh *et al*. No lipid class showing a significant systematic deviation except for SE, which showed slightly lower concentrations relative to this study, even after the application of RF. The comprehensive comparison of lipid species concentrations across all methods is summarized in Table S8.

### 3.5 Statistical analysis

The new optimized and validated RP-UHPLC/MS/MS method was applied to a clinical cohort consisting of healthy controls (without cancer history) and patients diagnosed with PDAC at various tumor stages. Detailed clinical characteristics of all samples are summarized in Table S3. While our previous studies focused on similar types of samples, those analyses relied either on the lipid class separation coupled to high-resolution MS [20,21] or on the direct infusion combined with low-resolution MS [19]. In contrast, the present method provides substantially higher structural information, enabling the analysis of 338 lipid species from 21 lipid subclasses using the positive ion mode, compared with 183 lipid species using our routine UHPSFC workflow and 128 lipid species with the HILIC based approach [28].

Lipid concentrations were determined in serum samples from 55 healthy volunteers (27 males and 28 females) and 54 PDAC patients (26 males and 28 females). The pooled serum sample, prepared from aliquots of all individual samples, was used as a system quality control (QC). The determined concentrations (Table S9) were evaluated using both univariate and multivariate statistical methods. The clear separation between healthy controls and PDAC patients was visible already from the unsupervised PCA projection (Figure 3A). Periodically injected QC samples show tight clustering, indicating the high quality and reproducibility of the acquired data. Supervised OPLS-DA analysis further confirmed a distinct separation of two cohorts (Figure 3B). These visualization approaches were applied separately to male and female cohorts (Figure S4), where the male subgroups remained well discriminated and the separation in females was less distinct. The most dysregulated lipids were identified using the S-plot generated from the OPLS-DA model considering all samples regardless of gender (Figure S5) and statistical significance of individual lipids is summarized in Table S10. Results of these univariate parameters were visualized using a volcano plot (Figure 3C), and heat map combined with clustering analysis (Figure 3D), highlighting the most significantly altered lipids across samples. Finally, network mapping was employed to illustrate the dysregulation of these lipids within their metabolic pathways, separately for sphingolipids (Figure 3E), phospholipids and less polar lipids (Figure S6).

**Figure 3.**
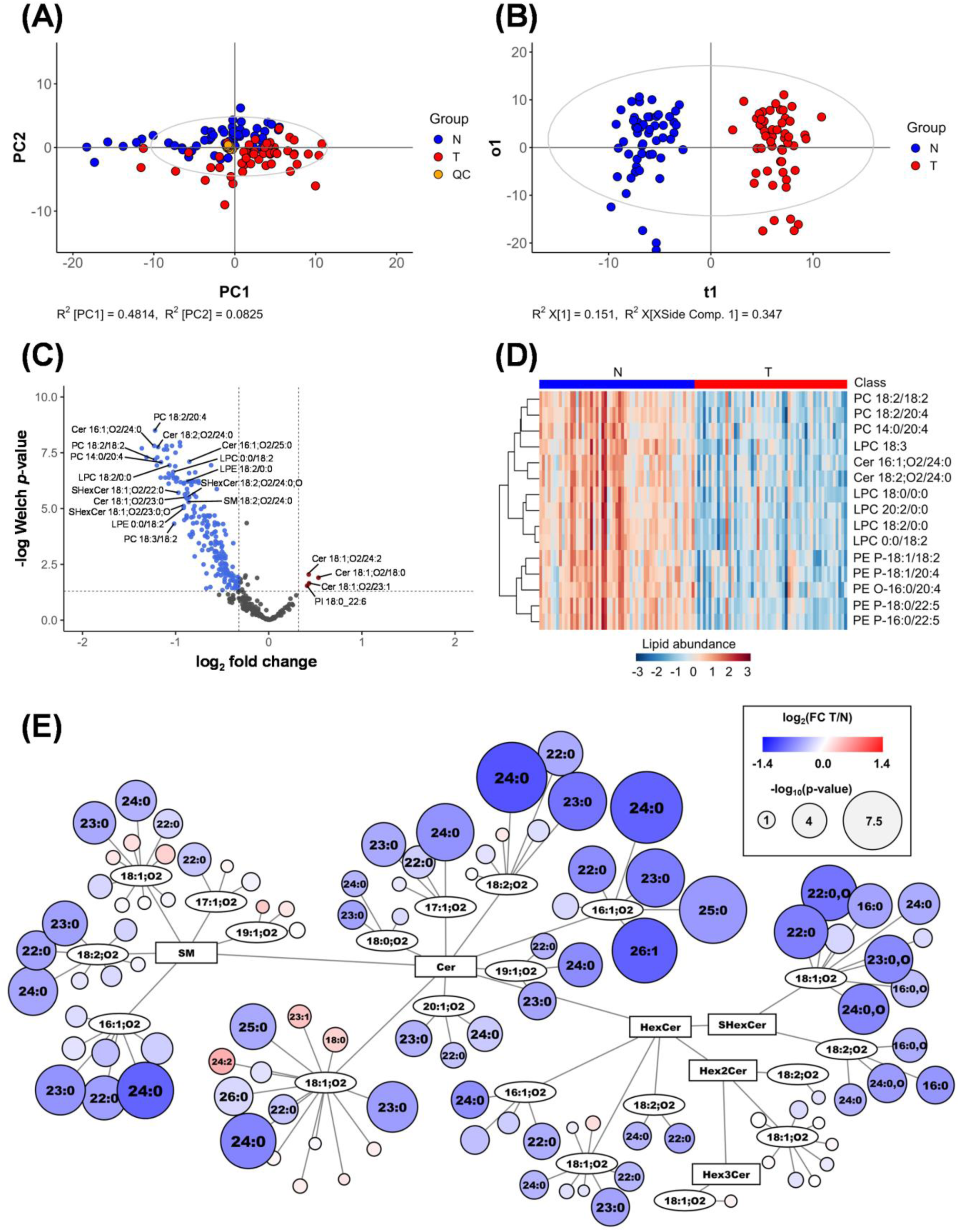
Results of the statistical analysis of PDAC patients (T, tumor) and healthy controls (N, normal). (A) Non-supervised PCA model. (B) Supervised OPLS-DA model. (C) Volcano plot highlighting the most upregulated (red) and downregulated (blue) lipids in PDAC. (D) Heat map of the most statistically significant lipid species. (E) Network map of sphingolipid species, where circle size reflects the *p*-value of individual lipids and red/blue color saturation represents the fold change (T/N).

### 3.6 Lipidomic alterations in PDAC

As already mentioned, the RP-UHPLC/MS/MS method not only covers a higher number of lipid subclasses compared to our routine approaches, but it also provides more detailed information on the lipid composition at the fatty acyl/alkyl level. The separation at the species level provides the sensitivity needed to distinguish small, but biologically important changes that would remain hidden using lipid class profiles. This level of detail is particularly critical in cancer research, as lipid dysregulation is often linked to certain species within a given class, reflecting specific enzymatic activities and metabolic or signaling pathways. This detailed resolution enabled us to identify the most statistically significant lipid subclasses, including SM, Cer, HexCer, SHexCer, PC, PE P-, LPC, and LPE, which is consistent with our previously published results based on the lipid class separation. In contrast, SE, DG, Hex2Cer, Hex3Cer, LPI, PI, and PE were found to be less significant.

The most important finding from previous studies was the downregulation of sphingolipids with compositions 39:1;O2, 40:1;O2, 41:1;O2, and 42:1;O2, together with the upregulation of 34:1;O2 and 36:1;O2 associated with the various types of cancer [11,18–21]. This trend highlights the importance of resolving the sphingoid base and *N*-acyl chain compositions, as the statistical relevance often follows the most abundant isomer (Figure 4A and Figure S7). In agreement with these reports, present results demonstrate that dysregulation of sphingolipids was largely determined by the composition of attached *N*-acyl chains (Figure 4B) rather than the sphingoid base (Figure 4C). A marked downregulation was observed for Cer, SM, HexCer, and SHexCer species (Figure S8) with linked very long chain saturated fatty acyls (VLCFA, ≥ C22), whereas Cer and SM species with shorter saturated chains (16:0 and 18:0) or with unsaturated chains (23:1, 24:1, and 24:2) showed slight upregulation. These patterns are clearly visualized on Figure 4D mapping *N*-acyl chain dependency for Cer and SM and for other sphingolipid subclasses on Figure S9. The specificity of this pattern is biochemically significant. The synthesis of VLCFA containing ceramides is predominantly catalyzed by ceramide synthase 2 (CerS2), while fatty acid desaturation is governed by enzymes such as stearoyl-CoA desaturase-1 (SCD1), whose upregulation is a known hallmark of many cancers [46–49]. The opposing trends we observe could therefore suggest a complex interplay between these distinct enzymatic pathways in PDAC. While a definitive elucidation of these mechanisms is beyond the scope of the present lipidomic study, our findings provide a clear, data-driven rationale for future investigations into the specific roles of ceramide synthases and desaturases in pancreatic cancer pathology. This level of structural information on sphingolipid dysregulation in cancer, which has not been previously reported, highlight the important role of RP-UHPLC/MS/MS in formulating biologically relevant hypotheses.

**Figure 4.**
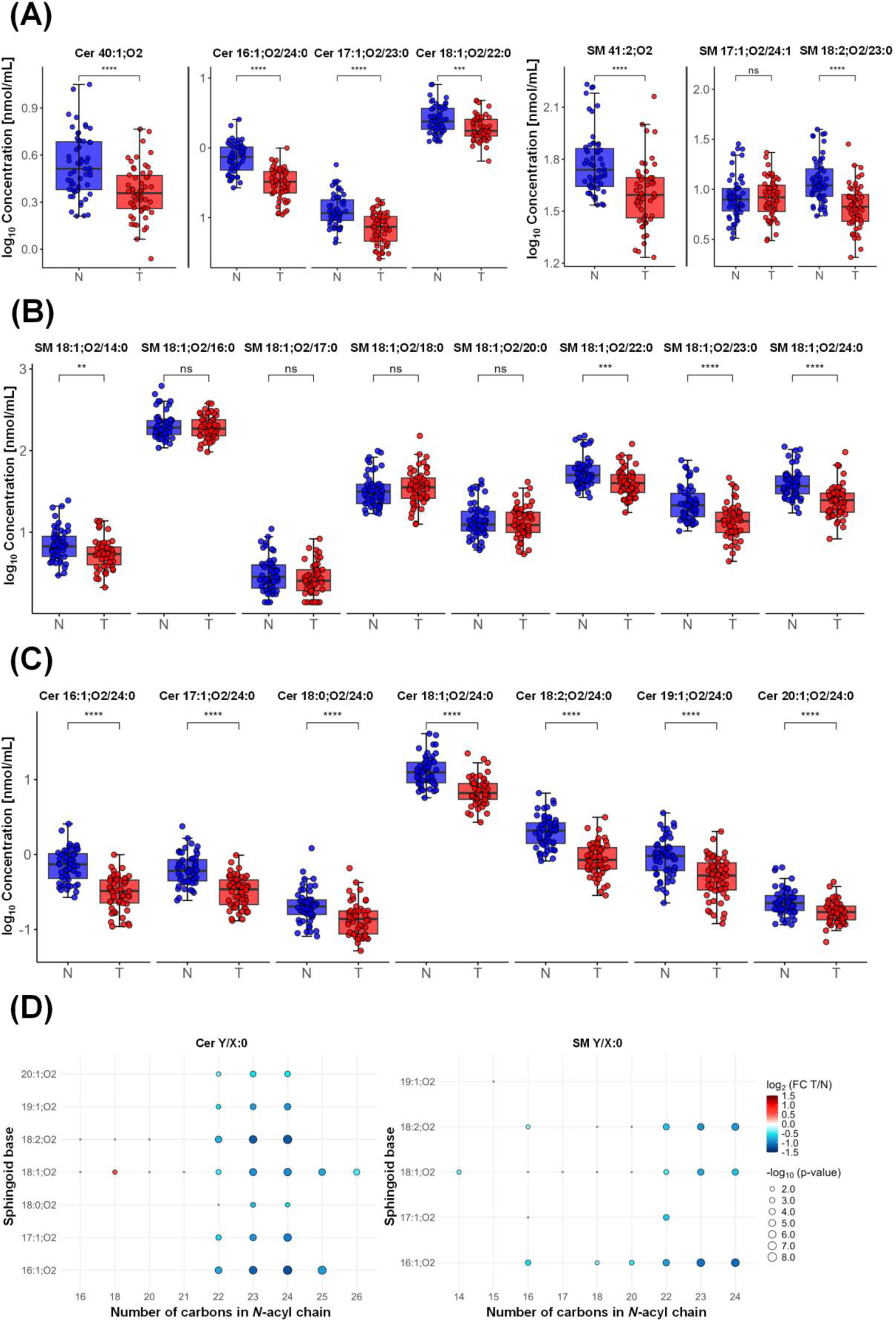
Statistical significance of selected ceramides and sphingomyelins between PDAC patients (T, tumor) and healthy controls (N, normal). (A) Comparison of lipid species level (sum of isomers) *vs.* fatty acyl level (individual isomers). (B) Statistical significance based on *N*-acyl chain composition. (C) Statistical significance based on sphingoid base composition. (D) Bubble plots showing the influence of various saturated *N*-acyl chain, where Y represents the sphingoid base and X denotes the number of carbons in the *N*-acyl chain.

Lysophospholipids were predominantly downregulated, with LPC 18:3, LPC 18:2, and LPE 18:2 identified as the most significant species. Importantly, both *sn-1* and *sn-2* isomers show comparable dysregulation, suggesting that the position of the fatty acyl chain may not play a major role in PDAC associated lipid alterations (Figure S10). Within phospholipid species, mainly PC, PC P-, PE P- and PE O-, consistent downregulation was observed for species containing fatty acyl 18:2 or 20:4 (e.g., PC 18:2/20:4, PC 18:2/18:2, PE P-18:0/18:2, and PE O-16:0/20:4). Phospholipids containing docosahexaenoic acid (FA 22:6) in the *sn-2* position (e.g., PI 18:0_22:6, PE 16:0/22:6, PE 18:0/22:6, and PE 18:1/22:6) were mildly upregulated. Notably, PE P- and PE O-subclasses were more strongly downregulated compared to PE, which was statistically non-significant and most likely reflects distinct metabolic pathways. The most significant changes were observed in plasmalogens containing polyunsaturated fatty acids, particularly 18:2, 20:3, 20:4, and 22:5. These findings suggest that alterations depend mainly on the fatty acyl composition at the *sn-2* position, rather than on the alkenyl chain. Box plots of the most statistically significant phospholipids including their ethers and plasmalogens are shown in Figure S11.

These dysregulations in phospholipids were further confirmed by GC/MS analysis (Figure S12), using a method specifically developed for the separation of FAMEs, prepared by alkaline hydrolysis of total lipid extracts. For this purpose, 30 samples from healthy volunteers and 30 samples from PDAC patients were analyzed (Table S11). Clear upregulation was observed for monounsaturated fatty acid (FA) 18:1 (9Z) and FA 18:1 (11Z) as well as for docosahexaenoic acid (FA 22:6). In contrast, the most strongly downregulated FA included FA 20:3 (5Z, 11Z, 14Z), FA 20:4 (8Z, 11Z, 14Z, 17Z), and FA 18:3 (6Z, 9Z, 12Z). FA 18:2 was also found among the downregulated species. S-plot and box plots of the most dysregulated lipids are shown in Figure S13 and Figure S14. These finding are consistent with RP-UHPLC/MS/MS data.

## Conclusions

In this study, we developed, optimized, and validated an RP-UHPLC/MS/MS method for quantitative lipidomic profiling in human serum. This method enables the identification of 455 and quantification of 381 lipid species across 22 subclasses, providing broader lipid coverage compared to our previously reported approaches using lipid class separation. The application of response factors significantly improved the accuracy of sterol ester quantification. Finally, the method was applied to a clinical cohort of PDAC patients and healthy controls, where univariate and multivariate statistical analyses demonstrated a great group separation. The most prominent differences were observed in sphingolipids and phospholipids, which is in agreement with previous reports. Sphingolipid alterations were primarily determined by *N*-acyl chain composition, with very long-chain fatty acids (≥ C22) showing marked downregulation, while shorter or unsaturated species were slightly upregulated. Phospholipid dysregulation was strongly dependent on fatty acyl composition, with plasmalogens containing polyunsaturated fatty acids being most significant. These results highlight the importance of resolving lipid composition at the fatty acyl level for biological interpretation and hypothesis generation. While analyses based on lipid classes can indicate general trends, only separation at the species level reveals, which specific molecular changes may underlie altered enzymatic activities or signaling pathways. The observed dysregulation of sphingolipids and phospholipids in cancer may results from a number of processes [50–52], and without complementary multiomics approaches, including transcriptomic, proteomic, and metabolomic data, precise biological interpretation and formulation of specific hypotheses remain limited.

## Data Availability

All data produced in the present study are available upon reasonable request to the authors.

## Supplementary data

**Appendix A:** Chromatograms of internal standards, calibration curves, dependencies of cholesteryl ester response factors, multivariate data analysis, network maps, box plots, and bubble plots. (PDF file)

**Appendix B:** Internal standard mixtures, clinical information, response factors of cholesteryl esters, MS settings and list of MRM transitions, identification list of lipid species and comparison with literature, method validation, quantitative analysis and comparison with literature, and statistical analysis. (Excel file)

## Acknowledgments

This work was supported by the ERC Adv grant No. 101095860 (European Research Council), and OP JAK grant No. CZ.02.01.01/00/22_008/0004644 (Ministry of Education, Youth and Sports, Czech Republic).

## Author contributions

**Zuzana Lásko**: Data curation, Investigation, Methodology, Visualization, Writing - original draft. **Ondřej Peterka**: Conceptualization, Investigation, Methodology, Writing – review & editing. **Robert Jirásko**: Conceptualization, Supervision, Writing – review & editing. **Anna Taylor**: Visualization, Writing – review & editing. **Tomáš Hájek**: Investigation, Methodology, Writing – review & editing. **Beatrice Mohelníková-Duchoňová**: Resources, Writing – review & editing. **Martin Loveček**: Resources, Writing – review & editing. **Bohuslav Melichar**: Resources, Writing

– review & editing. **Michal Holčapek**: Conceptualization, Funding acquisition, Project administration, Supervision, Writing – review & editing.

## Compliance with ethical standards

All volunteers signed informed consent, and the ethical committee approved blood collection.

## REFERENCES

[1] National Cancer Institute, Cancer Stat Facts: Pancreatic Cancer. https://seer.cancer.gov/statfacts/html/pancreas.html, 2025 (accessed 15 September 2025)

[2] C. Sun, A.H. Rosendahl, D. Ansari, R. Andersson, Proteome-based biomarkers in pancreatic cancer, World J. Gastroenterol. 17 (2011) 4845–4852. 10.3748/wjg.v17.i44.4845.

[3] M. Tian, Y.Z. Cui, G.H. Song, M.J. Zong, X.Y. Zhou, Y. Chen, J.X. Han, Proteomic analysis identifies MMP-9, DJ-1 and A1BG as overexpressed proteins in pancreatic juice from pancreatic ductal adenocarcinoma patients, BMC Cancer 8 (2008). 10.1186/1471-2407-8-241.

[4] S. Kato, K. Honda, Use of biomarkers and imaging for early detection of pancreatic cancer, Cancers (Basel) 12 (2020) 1965. 10.3390/cancers12071965.

[5] O. Peterka, R. Jirásko, M. Chocholoušková, L. Kuchař, D. Wolrab, R. Hájek, D. Vrána, O. Strouhal, B. Melichar, M. Holčapek, Lipidomic characterization of exosomes isolated from human plasma using various mass spectrometry techniques, Biochim. Biophys. Acta Mol. Cell Biol. Lipids 1865 (2020) 158634. 10.1016/j.bbalip.2020.158634.

[6] Y.H. Soung, S. Ford, V. Zhang, J. Chung, Exosomes in cancer diagnostics, Cancers (Basel) 9 (2017) 8. 10.3390/cancers9010008.

[7] S.V. Bratman, S.Y.C. Yang, M.A.J. Iafolla, Z. Liu, A.R. Hansen, P.L. Bedard, S. Lheureux, A. Spreafico, A.A. Razak, S. Shchegrova, M. Louie, P. Billings, B. Zimmermann, H. Sethi, A. Aleshin, D. Torti, K. Marsh, J. Eagles, I. Cirlan, Y. Hanna, D.L. Clouthier, S.C. Lien, P.S. Ohashi, W. Xu, L.L. Siu, T.J. Pugh, Personalized circulating tumor DNA analysis as a predictive biomarker in solid tumor patients treated with pembrolizumab, Nat. Cancer 1 (2020) 873–881. 10.1038/s43018-020-0096-5.

[8] E. Sánchez-Herrero, R. Serna-Blasco, L. Robado de Lope, V. González-Rumayor, A. Romero, M. Provencio, Circulating Tumor DNA as a Cancer Biomarker: An Overview of Biological Features and Factors That may Impact on ctDNA Analysis, Front. Oncol. 12 (2022). 10.3389/fonc.2022.943253.

[9] J. Hayes, P.P. Peruzzi, S. Lawler, MicroRNAs in cancer: Biomarkers, functions and therapy, Trends Mol. Med. 20 (2014) 460–469. 10.1016/j.molmed.2014.06.005.

[10] S. Cai, T. Pataillot-Meakin, A. Shibakawa, R. Ren, C.L. Bevan, S. Ladame, A.P. Ivanov, J.B. Edel, Single-molecule amplification-free multiplexed detection of circulating microRNA cancer biomarkers from serum, Nat. Commun. 12 (2021). 10.1038/s41467-021-23497-y.

[11] D. Wolrab, R. Jirásko, O. Peterka, J. Idkowiak, M. Chocholoušková, Z. Vaňková, K. Hořejší, I. Brabcová, D. Vrána, H. Študentová, B. Melichar, M. Holčapek, Plasma lipidomic profiles of kidney, breast and prostate cancer patients differ from healthy controls, Sci. Rep. 11 (2021). 10.1038/s41598-021-99586-1.

[12] M. Chocholoušková, R. Jirásko, D. Vrána, J. Gatěk, B. Melichar, M. Holčapek, Reversed phase UHPLC/ESI-MS determination of oxylipins in human plasma: a case study of female breast cancer, Anal. Bioanal. Chem. 411 (2019) 1239–1251. 10.1007/s00216-018-1556-y.

[13] E. Dorado, M.L. Doria, A. Nagelkerke, J.S. McKenzie, S. Maneta-Stavrakaki, T.E. Whittaker, J.K. Nicholson, R.C. Coombes, M.M. Stevens, Z. Takats, Extracellular vesicles as a promising source of lipid biomarkers for breast cancer detection in blood plasma, J. Extracell. Vesicles 13 (2024) e12419. 10.1002/jev2.12419.

[14] E. Cífková, R. Brumarová, M. Ovčačíková, D. Dobešová, K. Mičová, A. Kvasnička, Z. Vaňková, J. Šiller, L. Sákra, D. Friedecký, M. Holčapek, Lipidomic and metabolomic analysis reveals changes in biochemical pathways for non-small cell lung cancer tissues, Biochim. Biophys. Acta Mol. Cell Biol. Lipids 1867 (2022). 10.1016/j.bbalip.2021.159082.

[15] G. Wang, M. Qiu, X. Xing, J. Zhou, H. Yao, M. Li, R. Yin, Y. Hou, Y. Li, S. Pan, Y. Huang, F. Yang, F. Bai, H. Nie, S. Di, L. Guo, Z. Meng, J. Wang, Y. Yin, Lung cancer scRNA-seq and lipidomics reveal aberrant lipid metabolism for early-stage diagnosis, Sci. Transl. Med. 1 4 (2022). 10.1126/scitranslmed.abk2756.

[16] V. Caponigro, A.L. Tornesello, F. Merciai, D. La Gioia, E. Salviati, M.G. Basilicata, S. Musella, F. Izzo, A.S. Megna, L. Buonaguro, E. Sommella, F.M. Buonaguro, M.L. Tornesello, P. Campiglia, Integrated plasma metabolomics and lipidomics profiling highlights distinctive signature of hepatocellular carcinoma in HCV patients, J. Transl. Med. 21 (2023). 10.1186/s12967-023-04801-4.

[17] M. Masoodi, A. Gastaldelli, T. Hyötyläinen, E. Arretxe, C. Alonso, M. Gaggini, J. Brosnan, Q.M. Anstee, O. Millet, P. Ortiz, J.M. Mato, J.-F. Dufour, M. Orešič, Metabolomics and lipidomics in NAFLD: biomarkers and non-invasive diagnostic tests, Nat. Rev. Gastroenterol. Hepatol. 18 (2021) 835–856. 10.1038/s41575-021-00502-9.

[18] R. Jirásko, J. Idkowiak, D. Wolrab, A. Kvasnička, D. Friedecký, K. Polański, H. Študentová, V. Študent, B. Melichar, M. Holčapek, Altered Plasma, Urine, and Tissue Profiles of Sulfatides and Sphingomyelins in Patients with Renal Cell Carcinoma, Cancers (Basel) 14 (2022). 10.3390/cancers14194622.

[19] J. Idkowiak, R. Jirásko, D. Kolářová, J. Bártl, T. Hájek, M. Antonelli, Z. Vaňková, D. Wolrab, R. Hrstka, H. Študentová, B. Melichar, K. Pešková, M. Holčapek, Robust and high-throughput lipidomic quantitation of human blood samples using flow injection analysis with tandem mass spectrometry for clinical use, Anal. Bioanal. Chem. 415 (2023) 935–951. 10.1007/s00216-022-04490-w.

[20] O. Peterka, A. Maccelli, R. Jirásko, Z. Vaňková, J. Idkowiak, R. Hrstka, D. Wolrab, M. Holčapek, HILIC/MS quantitation of low-abundant phospholipids and sphingolipids in human plasma and serum: Dysregulation in pancreatic cancer, Anal. Chim. Acta 1288 (2024). 10.1016/j.aca.2023.342144.

[21] D. Wolrab, R. Jirásko, E. Cífková, M. Höring, D. Mei, M. Chocholoušková, O. Peterka, J. Idkowiak, T. Hrnčiarová, L. Kuchař, R. Ahrends, R. Brumarová, D. Friedecký, G. Vivo-Truyols, P. Škrha, J. Škrha, R. Kučera, B. Melichar, G. Liebisch, R. Burkhardt, M.R. Wenk, A. Cazenave-Gassiot, P. Karásek, I. Novotný, K. Greplová, R. Hrstka, M. Holčapek, Lipidomic profiling of human serum enables detection of pancreatic cancer, Nat. Commun. 13 (2022). 10.1038/s41467-021-27765-9.

[22] M.J. Conroy, R.M. Andrews, S. Andrews, L. Cockayne, E.A. Dennis, E. Fahy, C. Gaud, W.J. Griffiths, G. Jukes, M. Kolchin, K. Mendivelso, A.F. Lopez-Clavijo, C. Ready, S. Subramaniam, V.B. O’Donnell, LIPID MAPS: update to databases and tools for the lipidomics community, Nucleic Acids Res. 52 (2024) D1677–D1682. 10.1093/nar/gkad896.

[23] D. Wolrab, R. Jirásko, M. Chocholoušková, O. Peterka, M. Holčapek, Oncolipidomics: Mass spectrometric quantitation of lipids in cancer research, TrAC - Trends in Analytical Chemistry 120 (2019). 10.1016/j.trac.2019.04.012.

[24] R. Tabassum, J.T. Rämö, P. Ripatti, J.T. Koskela, M. Kurki, J. Karjalainen, P. Palta, S. Hassan, J. Nunez-Fontarnau, T.T.J. Kiiskinen, S. Söderlund, N. Matikainen, M.J. Gerl, M.A. Surma, C. Klose, N.O. Stitziel, H. Laivuori, A.S. Havulinna, S.K. Service, V. Salomaa, M. Pirinen, A. Jalanko, J. Kaprio, K. Donner, M. Kaunisto, N. Mars, A. Dada, A. Shcherban, A. Ganna, A. Lehisto, E. Kilpeläinen, G. Brein, G. Awaisa, J. Harju, K. Pärn, P.D.B. Parolo, R. Kajanne, S. Lemmelä, T.P. Sipilä, T. Sipilä, U. Lyhs, V. Llorens, T. Niiranen, K. Kristiansson, L. Männikkö, M.G. Jiménez, M. Perola, R. Wong, T. Kilpi, T. Hiekkalinna, E. Järvensivu, E. Kaiharju, H. Mattsson, M. Laukkanen, P. Laiho, S. Lähteenmäki, T. Sistonen, S. Soini, A. Ziemann, A. Lehtonen, A. Lertratanakul, B. Georgantas, B. Riley-Gillis, D. Quarless, F. Rahimov, G. Heap, H. Jacob, J. Waring, J.W. Davis, N. Smaoui, R. Popovic, S. Esmaeeli, J. Waring, A. Matakidou, B. Challis, D. Close, S. Petrovski, A. Karlsson, J. Schleutker, K. Pulkki, P. Virolainen, L. Kallio, A. Mannermaa, S. Heikkinen, V.M. Kosma, C.Y. Chen, H. Runz, J. Liu, P. Bronson, S. John, S. Lahdenperä, S. Eaton, W. Zhou, M. Hendolin, O. Tuovila, R. Pakkanen, J. Maranville, K. Usiskin, M. Hochfeld, R. Plenge, R. Yang, S. Biswas, S. Greenberg, E. Laakkonen, J. Kononen, J. Paloneva, U. Kujala, T. Kuopio, J. Laukkanen, E. Kangasniemi, K. Savinainen, R. Laaksonen, M. Arvas, J. Ritari, J. Partanen, K. Hyvärinen, T. Wahlfors, A. Peterson, D. Oh, D. Chang, E. Teng, E. Strauss, G. Kerchner, H. Chen, H. Chen, J. Schutzman, J. Michon, J. Hunkapiller, M. McCarthy, N. Bowers, T. Lu, T. Bhangale, D. Pulford, D. Waterworth, D. Kulkarni, F. Xu, J. Betts, J.E. Gordillo, J. Hoffman, K. Auro, L. McCarthy, S. Ghosh, M. Ehm, K. Pitkänen, T. Mäkelä, A. Loukola, H. Joensuu, J. Sinisalo, K. Eklund, L. Aaltonen, M. Färkkilä, O. Carpen, P. Kauppi, P. Tienari, T. Ollila, T. Tuomi, T. Meretoja, A. Pitkäranta, J. Turunen, K. Hannula-Jouppi, S. Pikkarainen, S. Seitsonen, M. Koskinen, A. Palomäki, J. Rinne, K. Metsärinne, K. Elenius, L. Pirilä, L. Koulu, M. Voutilainen, M. Juonala, S. Peltonen, V. Aaltonen, A. Loboda, A. Podgornaia, A. Chhibber, A. Chu, C. Fox, D. Diogo, E. Holzinger, J. Eicher, P. Gormley, V. Mehta, X. Wang, J. Kettunen, K. Pylkäs, M. Kalaoja, M. Karjalainen, R. Hinttala, R. Kaarteenaho, S. Vainio, T. Mantere, A. Remes, J. Huhtakangas, J. Junttila, K. Tasanen, L. Huilaja, M. Luodonpää, N. Hautala, P. Karihtala, S. Kauppila, T. Harju, T. Blomster, H. Soininen, I. Harvima, J. Pihlajamäki, K. Kaarniranta, M. Pelkonen, M. Laakso, M. Hiltunen, M. Kiviniemi, O. Kaipiainen-Seppänen, P. Auvinen, R. Kälviäinen, V. Julkunen, A. Malarstig, Å. Hedman, C. Marshall, C. Whelan, H. Lehtonen, J. Parkkinen, K. Linden, K. Kalpala, M. Miller, N. Bing, S. McDonough, X. Chen, X. Hu, Y. Wu, A. Auranen, A. Jussila, H. Uusitalo-Järvinen, H. Kankaanranta, H. Uusitalo, J. Peltola, M. Kähönen, P. Isomäki, T. Laitinen, T. Salmi, A. Muslin, C. Wang, C. Chatelain, E. Xu, F. Auge, K. Call, K. Klinger, M. Crohns, M. Gossel, K. Palin, M. Rivas, H. Siirtola, J.G. Tabuenca, M. Jauhiainen, M.J. Daly, N.B. Freimer, A. Palotie, M.R. Taskinen, K. Simons, S. Ripatti, Genetic architecture of human plasma lipidome and its link to cardiovascular disease, Nat. Commun. 10 (2019). 10.1038/s41467-019-11954-8.

[25] J.Y.H. Seah, W.S. Chew, F. Torta, C.M. Khoo, M.R. Wenk, D.R. Herr, H. Choi, E.S. Tai, R.M. van Dam, Plasma sphingolipids and risk of cardiovascular diseases: a large-scale lipidomic analysis, Metabolomics 16 (2020). 10.1007/s11306-020-01709-8.

[26] F. Carrillo, M. Ghirimoldi, G. Fortunato, N.P. Palomba, L. Ianiro, V. De Giorgis, S. Khoso, T. Giloni, S. Pietracupa, N. Modugno, E. Barberis, M. Manfredi, T. Esposito, Multiomics approach identifies dysregulated lipidomic and proteomic networks in Parkinson’s disease patients mutated in TMEM175, NPJ Parkinsons Dis. 11 (2025). 10.1038/s41531-024-00853-5.

[27] J. Hyuk Yoon, Y. Seo, Y. Suk Jo, S. Lee, E. Cho, A. Cazenave-Gassiot, Y.-S. Shin, M. Hee Moon, H. Joo An, M.R. Wenk, P.-G. Suh, Brain lipidomics: From functional landscape to clinical significance, Sci. Adv. 37 (2022). 10.1126/sciadv.adc9317

[28] D. Wolrab, M. Chocholoušková, R. Jirásko, O. Peterka, M. Holčapek, Validation of lipidomic analysis of human plasma and serum by supercritical fluid chromatography–mass spectrometry and hydrophilic interaction liquid chromatography–mass spectrometry, Anal. Bioanal. Chem. 412 (2020) 2375–2388. 10.1007/s00216-020-02473-3.

[29] M. Chocholoušková, F. Torta, Fast and comprehensive lipidomic analysis using supercritical fluid chromatography coupled with low and high resolution mass spectrometry, J. Chromatogr. A 1745 (2025). 10.1016/j.chroma.2025.465742.

[30] J.W. Lee, S. Nishiumi, M. Yoshida, E. Fukusaki, T. Bamba, Simultaneous profiling of polar lipids by supercritical fluid chromatography/tandem mass spectrometry with methylation, J. Chromatogr. A 1279 (2013) 98–107. 10.1016/j.chroma.2013.01.020.

[31] M. Ovčačíková, M. Lísa, E. Cífková, M. Holčapek, Retention behavior of lipids in reversed-phase ultrahigh-performance liquid chromatography–electrospray ionization mass spectrometry, J. Chromatogr. A 1450 (2016) 76–85. 10.1016/J.CHROMA.2016.04.082.

[32] Z. Vaňková, O. Peterka, M. Chocholoušková, D. Wolrab, R. Jirásko, M. Holčapek, Retention dependences support highly confident identification of lipid species in human plasma by reversed-phase UHPLC/MS, Anal. Bioanal. Chem. 414 (2022) 319–331. 10.1007/s00216-021-03492-4.

[33] K. Huynh, C.K. Barlow, K.S. Jayawardana, J.M. Weir, N.A. Mellett, M. Cinel, D.J. Magliano, J.E. Shaw, B.G. Drew, P.J. Meikle, High-Throughput Plasma Lipidomics: Detailed Mapping of the Associations with Cardiometabolic Risk Factors, Cell Chem. Biol. 26 (2019) 71–84. 10.1016/j.chembiol.2018.10.008.

[34] O. Peterka, Y. Kadyrbekova, R. Jirásko, Z. Lásko, B. Melichar, M. Holčapek, Novel Charge-Switch Derivatization Method Using 3-(Chlorosulfonyl)benzoic Acid for Sensitive RP-UHPLC/MS/MS Analysis of Acylglycerols, Sterols, and Prenols, Anal. Chem. 97 (2025) 7157–7164. 10.1021/acs.analchem.4c06496.

[35] O. Peterka, R. Jirásko, Z. Vaňková, M. Chocholoušková, D. Wolrab, J. Kulhánek, F. Bureš, M. Holčapek, Simple and Reproducible Derivatization with Benzoyl Chloride: Improvement of Sensitivity for Multiple Lipid Classes in RP-UHPLC/MS, Anal. Chem. 93 (2021) 13835–13843. 10.1021/acs.analchem.1c02463.

[36] K.J. Adams, B. Pratt, N. Bose, L.G. Dubois, L. St. John-Williams, K.M. Perrott, K. Ky, P. Kapahi, V. Sharma, M.J. Maccoss, M.A. Moseley, C.A. Colton, B.X. Maclean, B. Schilling, J.W. Thompson, Skyline for Small Molecules: A Unifying Software Package for Quantitative Metabolomics, J. Proteome Res. 19 (2020) 1447–1458. 10.1021/acs.jproteome.9b00640.

[37] G. Liebisch, E. Fahy, J. Aoki, E.A. Dennis, T. Durand, C.S. Ejsing, M. Fedorova, I. Feussner, W.J. Griffiths, H. Köfeler, A.H. Merrill, R.C. Murphy, V.B. O’Donnell, O. Oskolkova, S. Subramaniam, M.J.O. Wakelam, F. Spener, Update on LIPID MAPS classification, nomenclature, and shorthand notation for MS-derived lipid structures, J. Lipid Res. 61 (2020) 1539–1555. 10.1194/jlr.S120001025.

[38] Z. Lásko, T. Hájek, R. Jirásko, O. Peterka, P. Šimek, P.J. Schoenmakers, M. Holčapek, Four-Dimensional Lipidomic Analysis Using Comprehensive Online UHPLC × UHPSFC/Tandem Mass Spectrometry, Anal. Chem. 96 (2024) 19439–19446. 10.1021/acs.analchem.4c03946.

[39] R. Mandal, J. Zheng, L. Zhang, E. Oler, M.A. LeVatte, M. Berjanskii, M. Lipfert, J. Han, C.H. Borchers, D.S. Wishart, Comprehensive, Quantitative Analysis of SRM 1950: the NIST Human Plasma Reference Material, Anal. Chem. 97 (2025) 667–675. 10.1021/acs.analchem.4c05018.

[40] M. Ghorasaini, Y. Mohammed, J. Adamski, L. Bettcher, J.A. Bowden, M. Cabruja, K. Contrepois, M. Ellenberger, B. Gajera, M. Haid, D. Hornburg, C. Hunter, C.M. Jones, T. Klein, O. Mayboroda, M. Mirzaian, R. Moaddel, L. Ferrucci, J. Lovett, K. Nazir, M. Pearson, B.K. Ubhi, D. Raftery, F. Riols, R. Sayers, E.J.G. Sijbrands, M.P. Snyder, B. Su, V. Velagapudi, K.J. Williams, Y.B. De Rijke, M. Giera, Cross-Laboratory Standardization of Preclinical Lipidomics Using Differential Mobility Spectrometry and Multiple Reaction Monitoring, Anal. Chem. 93 (2021) 16369–16378. 10.1021/acs.analchem.1c02826.

[41] O. Quehenberger, A.M. Armando, A.H. Brown, S.B. Milne, D.S. Myers, A.H. Merrill, S. Bandyopadhyay, K.N. Jones, S. Kelly, R.L. Shaner, C.M. Sullards, E. Wang, R.C. Murphy, R.M. Barkley, T.J. Leiker, C.R.H. Raetz, Z. Guan, G.M. Laird, D.A. Six, D.W. Russell, J.G. McDonald, S. Subramaniam, E. Fahy, E.A. Dennis, Lipidomics reveals a remarkable diversity of lipids in human plasma1, J. Lipid Res. 51 (2010) 3299–3305. 10.1194/jlr.M009449.

[42] J.A. Bowden, A. Heckert, C.Z. Ulmer, C.M. Jones, J.P. Koelmel, L. Abdullah, L. Ahonen, Y. Alnouti, A.M. Armando, J.M. Asara, T. Bamba, J.R. Barr, J. Bergquist, C.H. Borchers, J. Brandsma, S.B. Breitkopf, T. Cajka, A. Cazenave-Gassiot, A. Checa, M.A. Cinel, R.A. Colas, S. Cremers, E.A. Dennis, J.E. Evans, A. Fauland, O. Fiehn, M.S. Gardner, T.J. Garrett, K.H. Gotlinger, J. Han, Y. Huang, A.H. Neo, T. Hyötyläinen, Y. Izumi, H. Jiang, H. Jiang, J. Jiang, M. Kachman, R. Kiyonami, K. Klavins, C. Klose, H.C. Köfeler, J. Kolmert, T. Koal, G. Koster, Z. Kuklenyik, I.J. Kurland, M. Leadley, K. Lin, K.R. Maddipati, D. McDougall, P.J. Meikle, N.A. Mellett, C. Monnin, M.A. Moseley, R. Nandakumar, M. Oresic, R. Patterson, D. Peake, J.S. Pierce, M. Post, A.D. Postle, R. Pugh, Y. Qiu, O. Quehenberger, P. Ramrup, J. Rees, B. Rembiesa, D. Reynaud, M.R. Roth, S. Sales, K. Schuhmann, M.L. Schwartzman, C.N. Serhan, A. Shevchenko, S.E. Somerville, L. St John-Williams, M.A. Surma, H. Takeda, R. Thakare, J.W. Thompson, F. Torta, A. Triebl, M. Trötzmüller, S.J.K. Ubhayasekera, D. Vuckovic, J.M. Weir, R. Welti, M.R. Wenk, C.E. Wheelock, L. Yao, M. Yuan, X.H. Zhao, S. Zhou, Harmonizing lipidomics: NIST interlaboratory comparison exercise for lipidomics using SRM 1950-Metabolites in frozen human plasma, J. Lipid Res. 58 (2017) 2275–2288. 10.1194/jlr.m079012.

[43] M. Holčapek, M. Lísa, P. Jandera, N. Kabátová, Quantitation of triacylglycerols in plant oils using HPLC with APCI-MS, evaporative light-scattering, and UV detection, J. Sep. Sci. 28 (2005) 1315–1333. 10.1002/jssc.200500088.

[44] M. Höring, C.S. Ejsing, M. Hermansson, G. Liebisch, Quantification of Cholesterol and Cholesteryl Ester by Direct Flow Injection High-Resolution Fourier Transform Mass Spectrometry Utilizing Species-Specific Response Factors, Anal. Chem. 91 (2019) 3459–3466. 10.1021/acs.analchem.8b05013.

[45] G. Liebisch, R. Ahrends, M. Arita, M. Arita, J.A. Bowden, C.S. Ejsing, W.J. Griffiths, M. Holčapek, H. Köfeler, T.W. Mitchell, M.R. Wenk, K. Ekroos, Lipidomics needs more standardization, Nat. Metab. 1 (2019) 745–747. 10.1038/s42255-019-0094-z.

[46] T. Pani, K. Rajput, A. Kar, H. Sharma, R. Basak, N. Medatwal, S. Saha, G. Dev, S. Kumar, S. Gupta, A. Mukhopadhyay, D. Malakar, T.K. Maiti, A.G. Arimbasseri, S.V.S. Deo, R.D. Sharma, A. Bajaj, U. Dasgupta, Alternative splicing of ceramide synthase 2 alters levels of specific ceramides and modulates cancer cell proliferation and migration in Luminal B breast cancer subtype, Cell Death Dis. 12 (2021). 10.1038/s41419-021-03436-x.

[47] S.D. Spassieva, T.D. Mullen, D.M. Townsend, L.M. Obeid, Disruption of ceramide synthesis by CerS2 down-regulation leads to autophagy and the unfolded protein response, Biochem. J. 424 (2009) 273–283. 10.1042/BJ20090699.

[48] U. Sen, C. Coleman, T. Sen, Stearoyl coenzyme A desaturase-1: multitasker in cancer, metabolism, and ferroptosis, Trends Cancer 9 (2023) 480–489. 10.1016/j.trecan.2023.03.003.

[49] Z. Tracz-Gaszewska, P. Dobrzyn, Stearoyl-CoA desaturase 1 as a therapeutic target for the treatment of cancer, Cancers (Basel) 11 (2019). 10.3390/cancers11070948.

[50] T. Kolter, K. Sandhoff, Sphingolipid metabolism diseases, BBA 1758 (2006) 2057–2079. 10.1016/j.bbamem.2006.05.027.

[51] D. Delmas, A. Mialhe, A.K. Cotte, J.L. Connat, F. Bouyer, F. Hermetet, V. Aires, Lipid metabolism in cancer: Exploring phospholipids as potential biomarkers, Biomedicine and Pharmacotherapy 187 (2025). 10.1016/j.biopha.2025.118095.

[52] R.Z. Li, X.R. Wang, J. Wang, C. Xie, X.X. Wang, H.D. Pan, W.Y. Meng, T.L. Liang, J.X. Li, P.Y. Yan, Q.B. Wu, L. Liu, X.J. Yao, E.L.H. Leung, The key role of sphingolipid metabolism in cancer: New therapeutic targets, diagnostic and prognostic values, and anti-tumor immunotherapy resistance, Front. Oncol. 12 (2022). 10.3389/fonc.2022.941643.

