## Supplementary materials for "RP-UHPLC/MS/MS Provides Enhanced Lipidomic Profiling of Human Serum in Pancreatic Cancer"

**Figure S1:** Chromatogram of the IS-mix used for lipid quantitation in human serum, measured by RP-UHPLC/MS/MS in the positive ion mode.

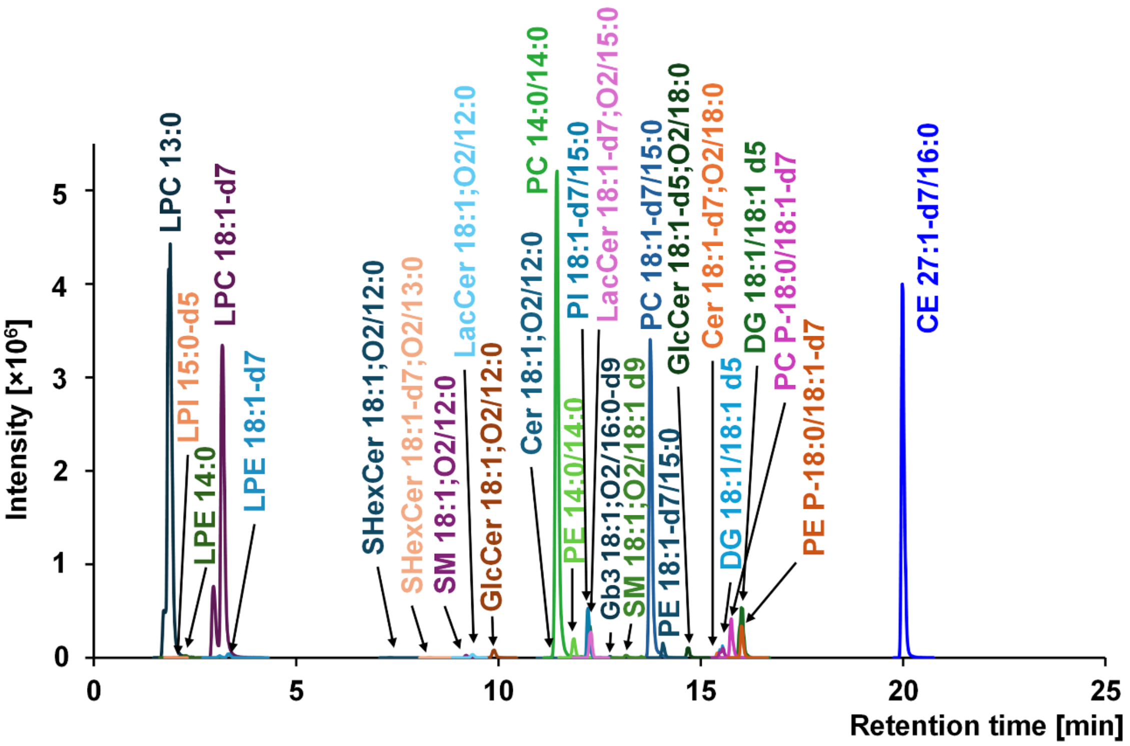

**Figure S2:** Calibration curves of internal standards spiked before the extraction. The data represent the mean values of three independent experiments. Internal standards used for the quantitative analysis are highlighted in blue. Regression equations and coefficients of determination ($R^2$) are provided in Table S6.

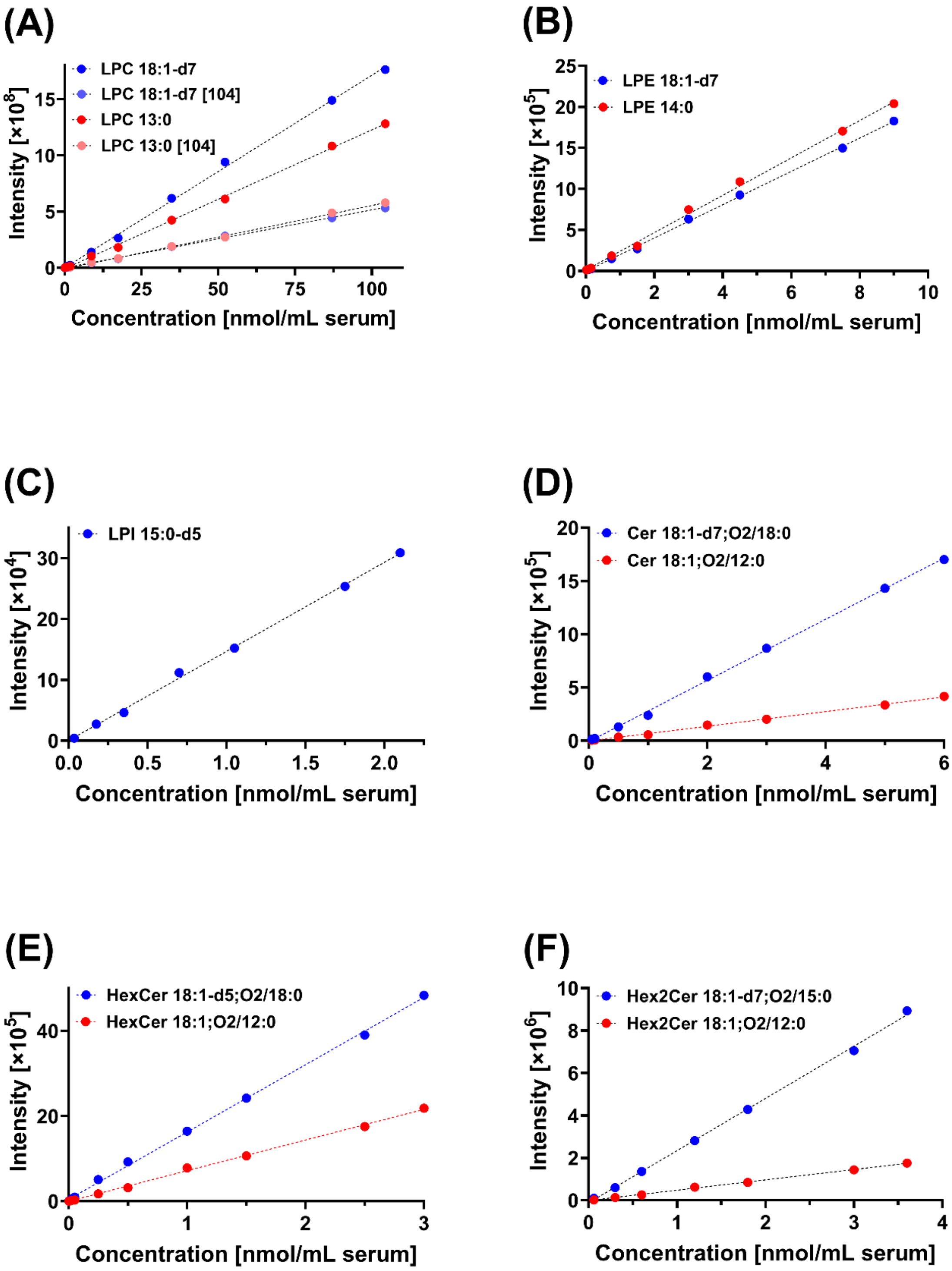

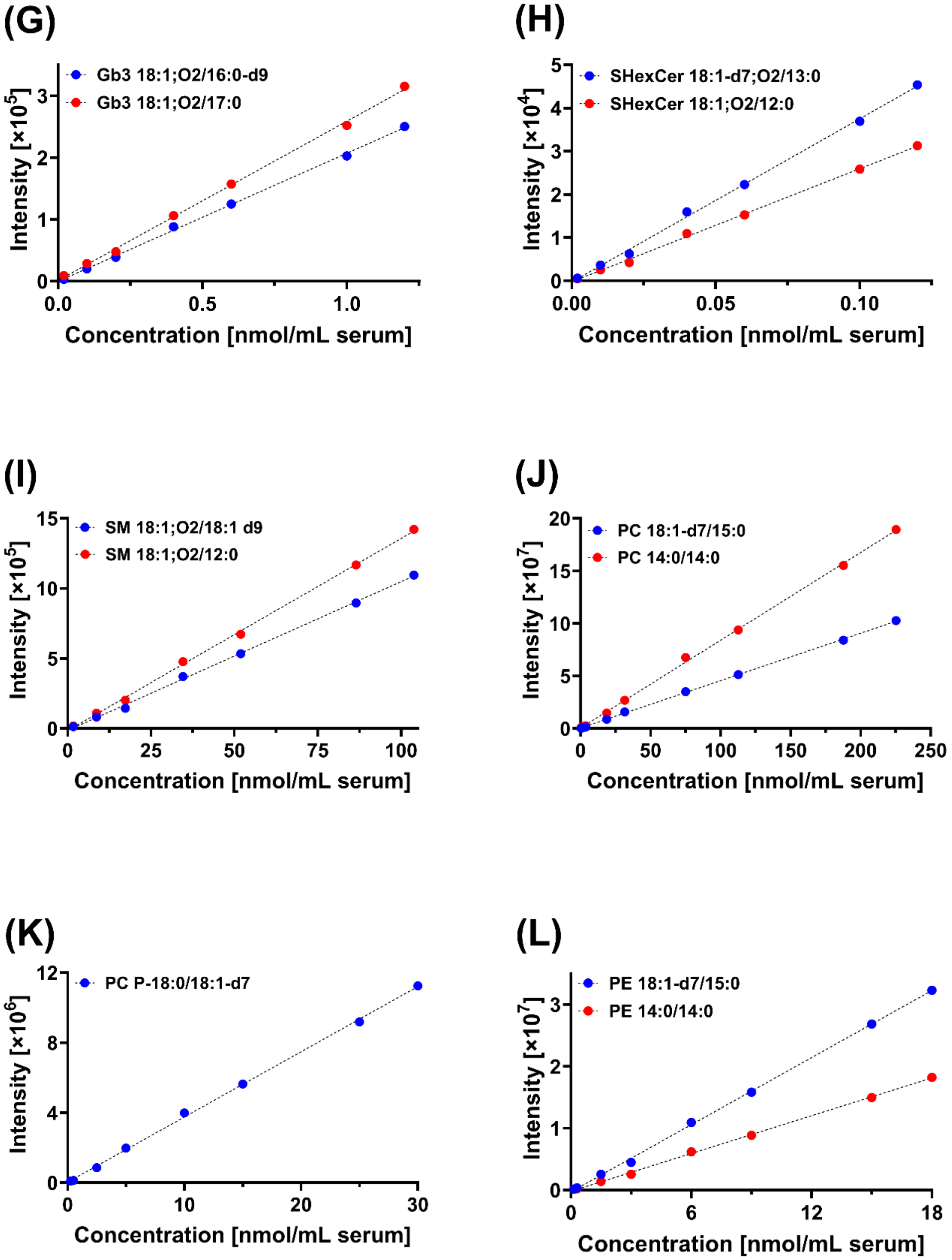
(G)
Gb3 18:1;O2/16:0-d9
Gb3 18:1;O2/17:0
Intensity [×10^5]
0
1
2
3
0.0
0.5
1.0
Concentration [nmol/mL serum]
(H)
SHexCer 18:1-d7;O2/13:0
SHexCer 18:1;O2/12:0
Intensity [×10^4]
0
1
2
3
4
5
0.00
0.05
0.10
Concentration [nmol/mL serum]
(I)
SM 18:1;O2/18:1 d9
SM 18:1;O2/12:0
Intensity [×10^5]
0
5
10
15
0
25
50
75
100
Concentration [nmol/mL serum]
(J)
PC 18:1-d7/15:0
PC 14:0/14:0
Intensity [×10^7]
0
5
10
15
20
0
50
100
150
200
250
Concentration [nmol/mL serum]
(K)
PC P-18:0/18:1-d7
Intensity [×10^6]
0
4
8
12
0
10
20
30
Concentration [nmol/mL serum]
(L)
PE 18:1-d7/15:0
PE 14:0/14:0
Intensity [×10^7]
0
1
2
3
0
6
12
18
Concentration [nmol/mL serum]

**(M)**

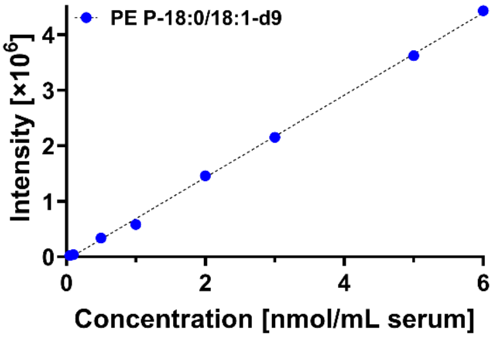
PE P-18:0/18:1-d9
Intensity [×10^6]
0
1
2
3
4
0
2
4
6
Concentration [nmol/mL serum]

**(N)**

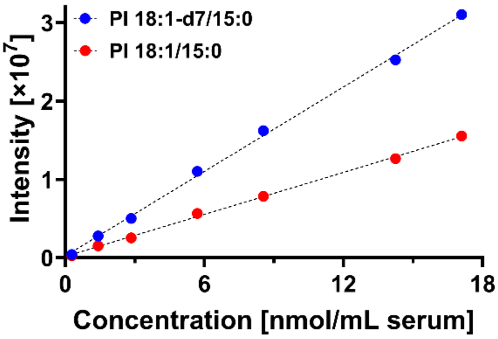
PI 18:1-d7/15:0
PI 18:1/15:0
Intensity [×10^7]
0
1
2
3
0
6
12
18
Concentration [nmol/mL serum]

**(O)**

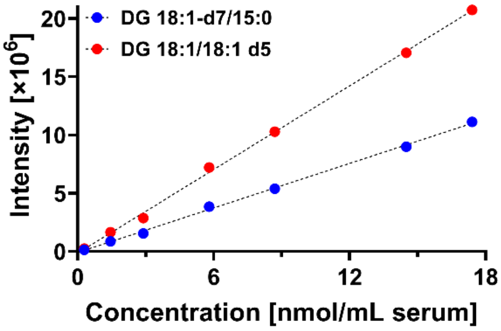
DG 18:1-d7/15:0
DG 18:1/18:1 d5
Intensity [×10^6]
0
5
10
15
20
0
6
12
18
Concentration [nmol/mL serum]

**(P)**

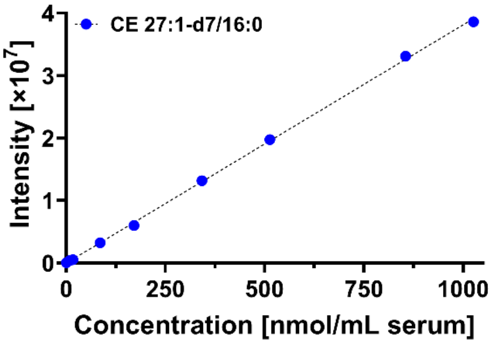
CE 27:1-d7/16:0
Intensity [×10^7]
0
1
2
3
4
0
250
500
750
1000
Concentration [nmol/mL serum]

**(Q)**

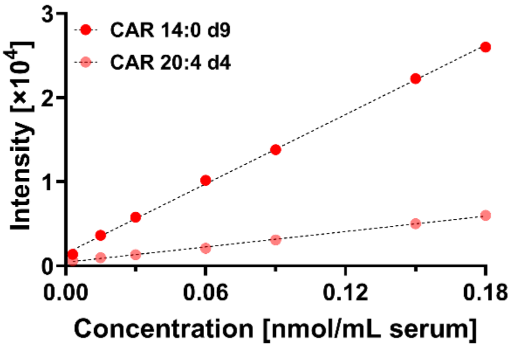
CAR 14:0 d9
CAR 20:4 d4
Intensity [×10^4]
0
1
2
3
0.00
0.06
0.12
0.18
Concentration [nmol/mL serum]

**Figure S3:** Dependences of response factors on fatty acyl chain length for (A) saturated and (B) monounsaturated cholesteryl esters, and on the number of double bonds for (C) C18 and (D) C20 fatty acyl chains of cholesteryl esters.

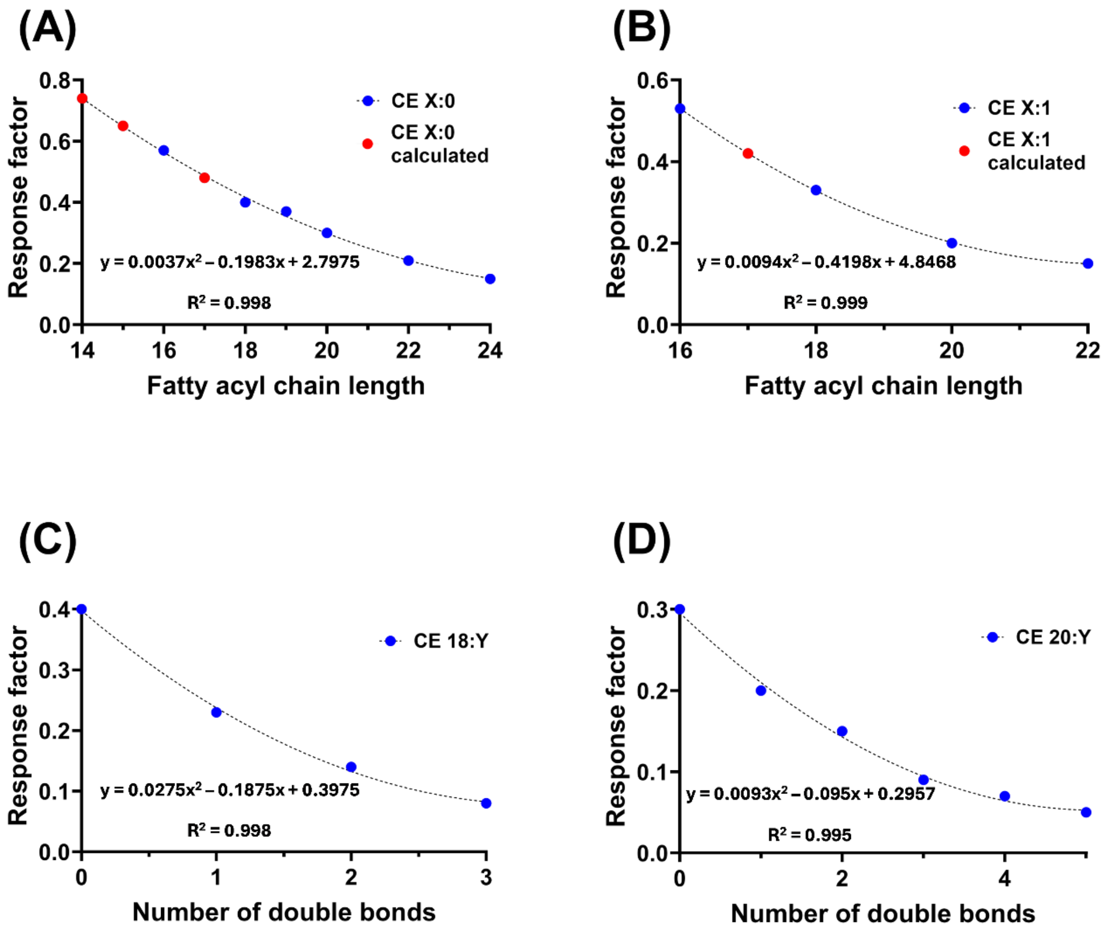

**Figure S4:** Lipidomic profiling results for serum samples of healthy controls and PDAC patients, including a non-supervised PCA model for females (A) and males (B), and supervised OPLS-DA model for females (C) and males (D).

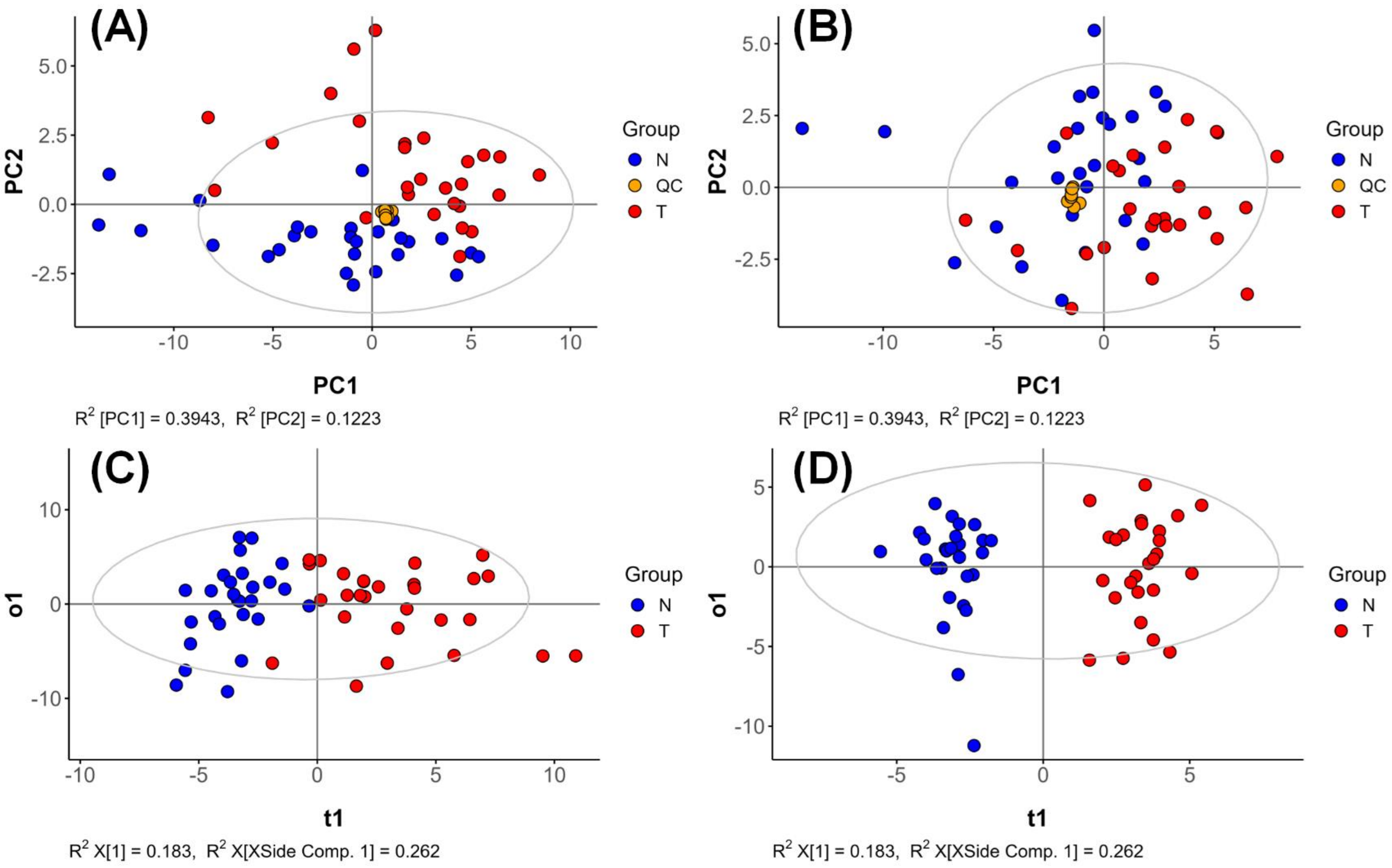

**Figure S5:** S-plot generated from OPLS-DA showing the most upregulated (red) and the most downregulated (blue) lipids in PDAC.

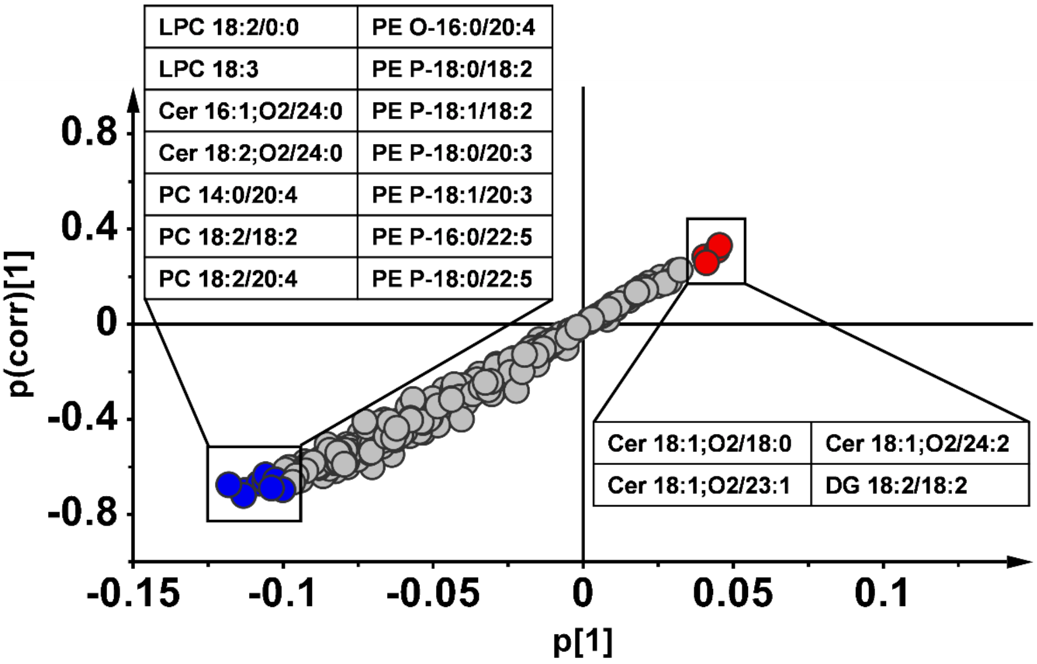

**Figure S6:** Network mapping of lipid species for (A) phospholipids and (B) diacylglycerols and sterol esters. Circles represent the detected lipid species, with circle size indicating statistical significance (*p*-value) and color intensity (red/blue) representing the degree of up- or down-regulation according to the fold change.

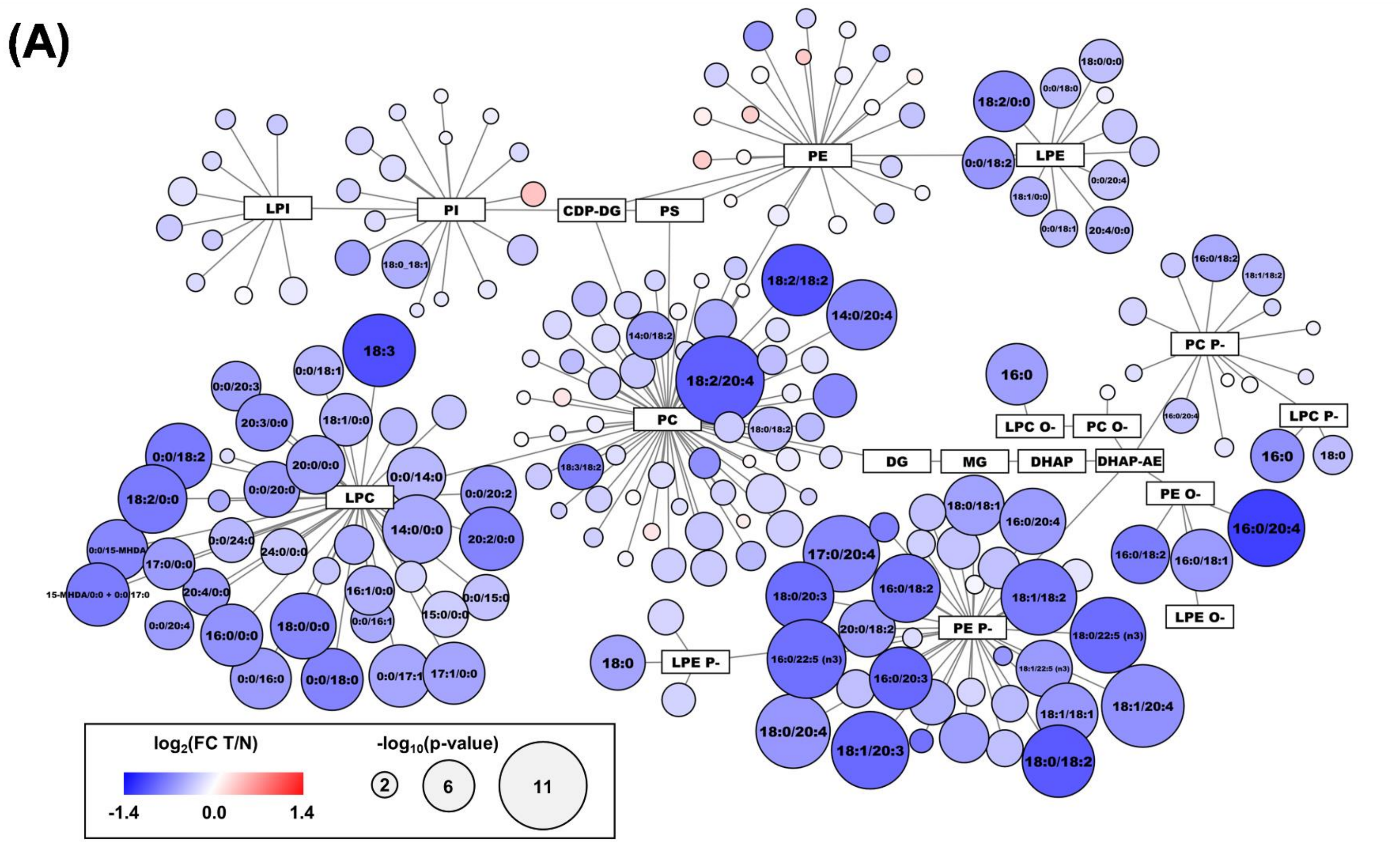

(B)

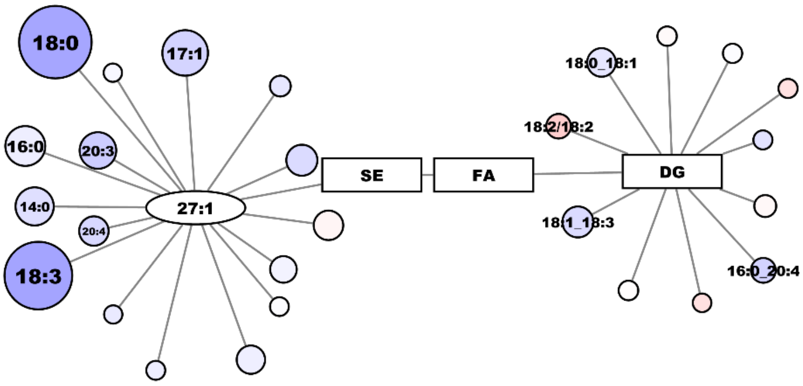
18:0
17:1
16:0
20:3
14:0
20:4
18:3
27:1
SE
FA
DG
18:0_18:1
18:2/18:2
18:1_18:3
16:0_20:4

**Figure S7:** Statistical comparison between lipid species level and fatty acyl level for (A) sphingomyelins and (B) ceramides. Significances were determined using the Mann–Whitney U test with the following thresholds: ns ($p > 0.05$), * ($p \leq 0.05$), ** ($p \leq 0.01$), *** ($p \leq 0.001$), and **** ($p \leq 0.0001$). N... healthy controls (blue); T... PDAC patients (red).

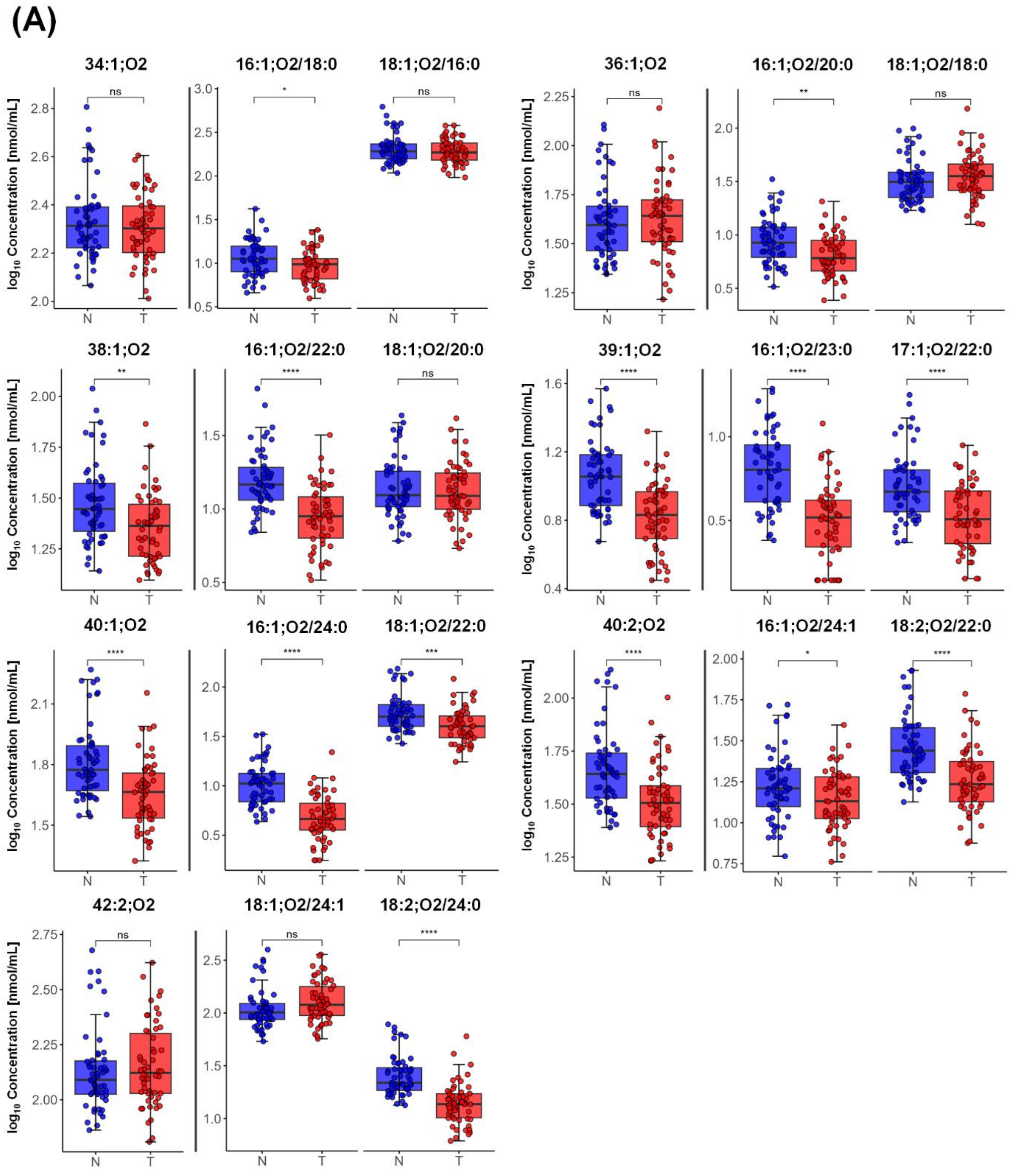

(B)

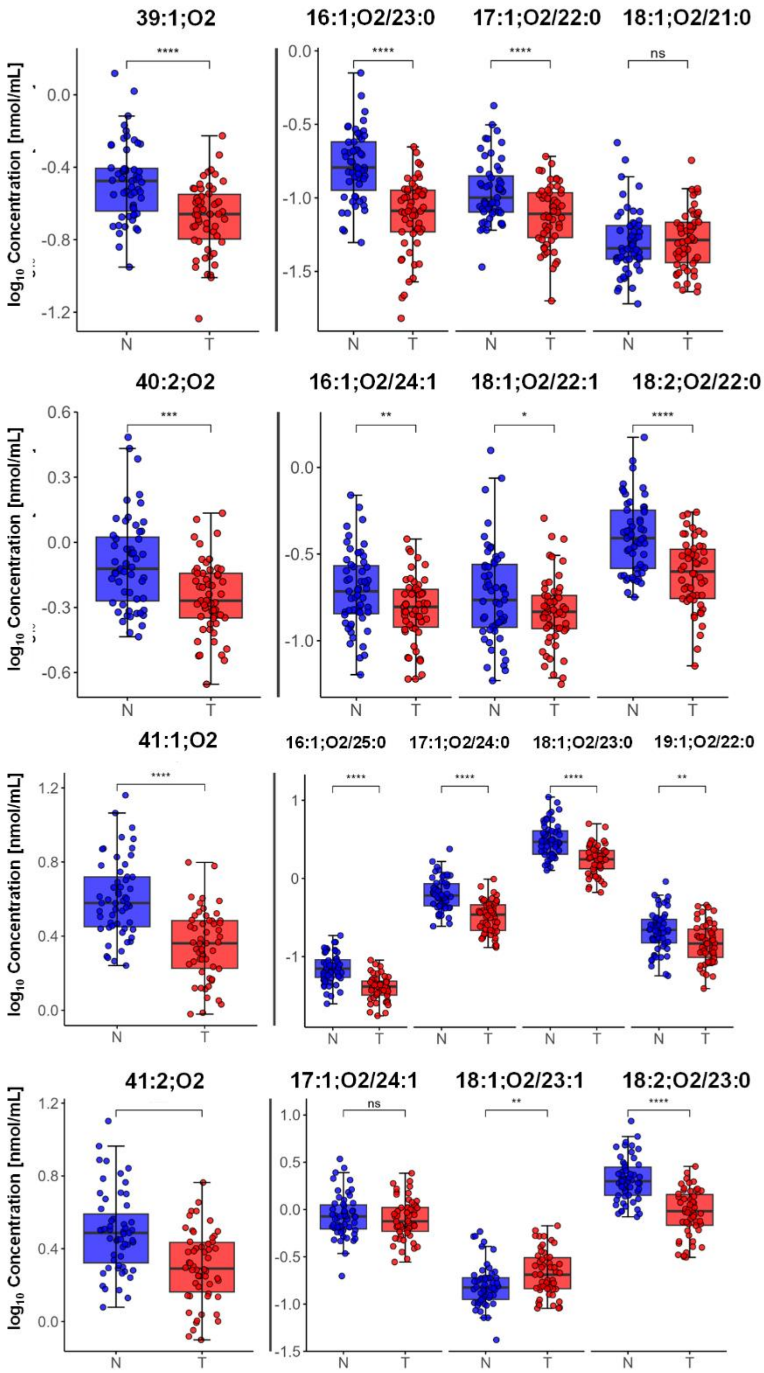
39:1;O2
16:1;O2/23:0
17:1;O2/22:0
18:1;O2/21:0
40:2;O2
16:1;O2/24:1
18:1;O2/22:1
18:2;O2/22:0
41:1;O2
16:1;O2/25:0
17:1;O2/24:0
18:1;O2/23:0
19:1;O2/22:0
41:2;O2
17:1;O2/24:1
18:1;O2/23:1
18:2;O2/23:0
log10 Concentration [nmol/mL]
N
T
****
***
**
*
ns

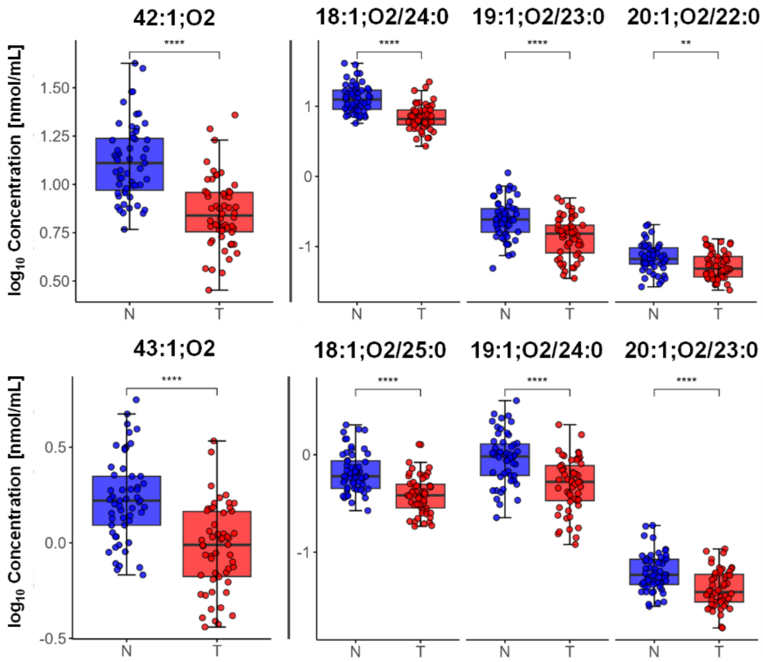

42:1;O2
18:1;O2/24:0
19:1;O2/23:0
20:1;O2/22:0
43:1;O2
18:1;O2/25:0
19:1;O2/24:0
20:1;O2/23:0
log10 Concentration [nmol/mL]
****
**
N
T

**Figure S8:** Statistical comparison of sphingolipids with 18:1;O2/24:0 fatty acyl composition. Significances were determined using the Mann–Whitney U test with the following thresholds: ns ($p > 0.05$), * ($p \leq 0.05$), ** ($p \leq 0.01$), *** ($p \leq 0.001$), **** ($p \leq 0.0001$). N... healthy controls (blue); T... PDAC patients (red).

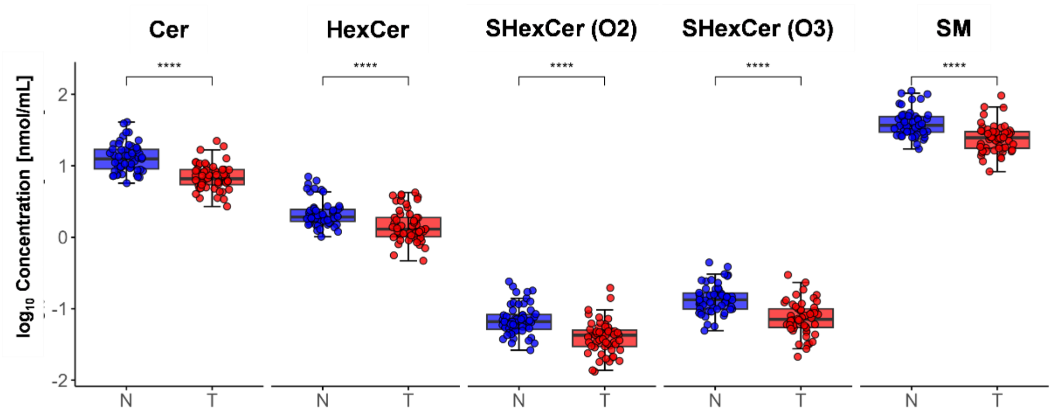

**Figure S9:** Bubble fatty acyl chain plots for (A) sphingomyelins, (B) ceramides, (C) hexosylceramides, and (D) sulfatides, illustrating their statistical significance in PDAC.

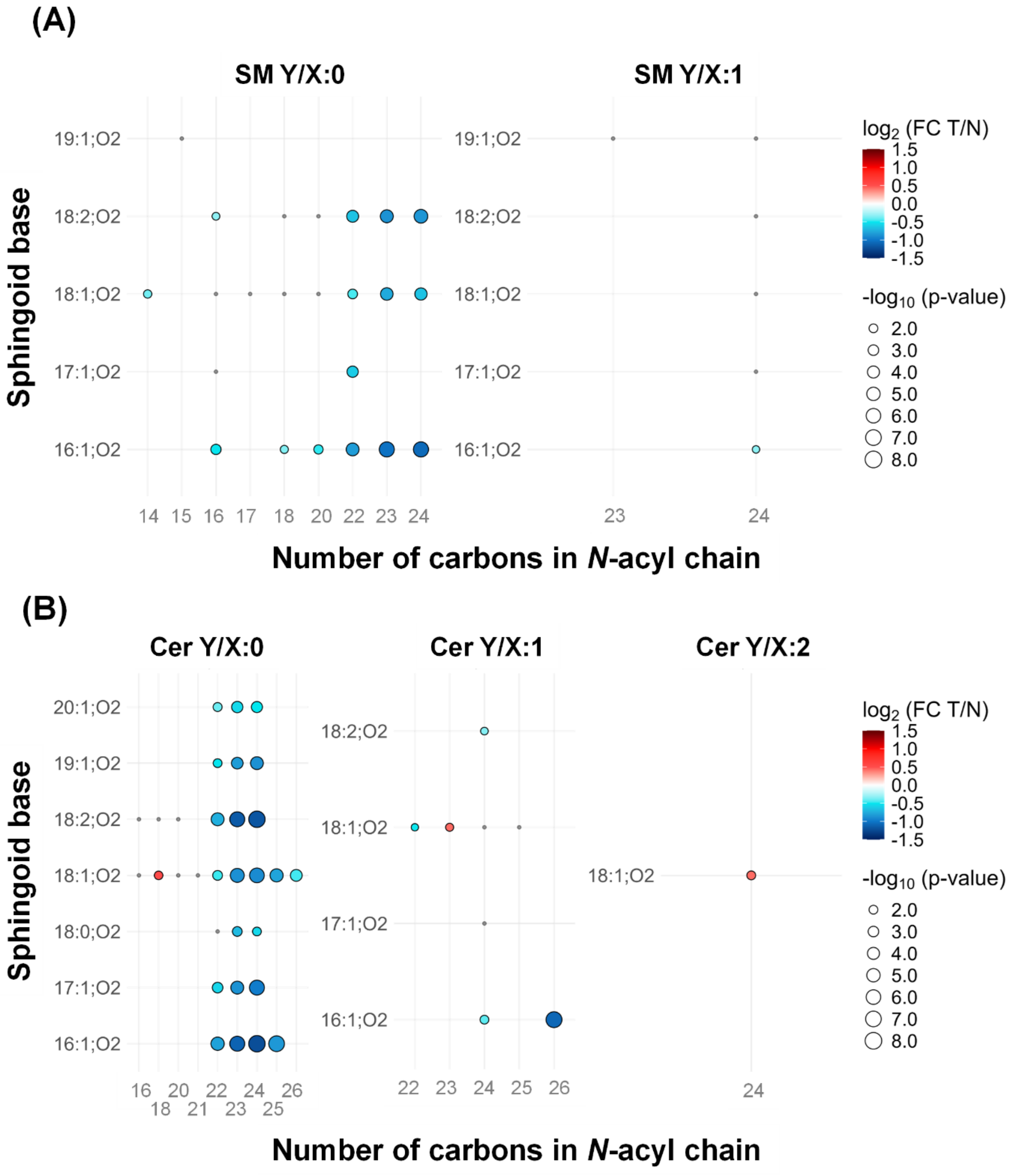

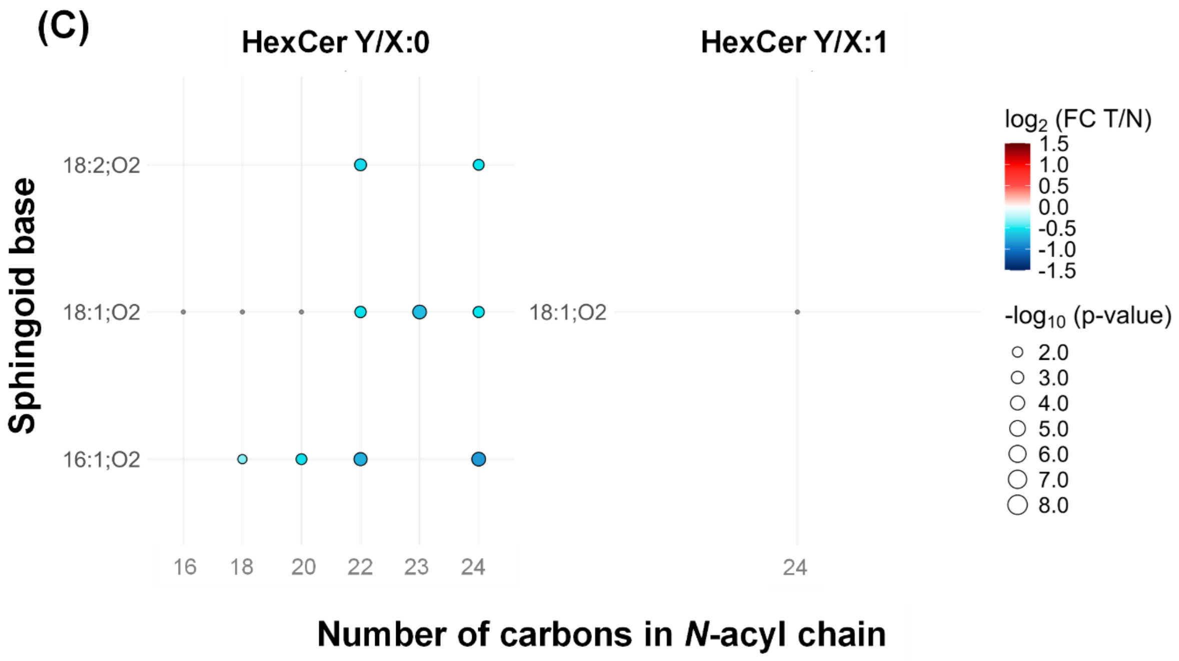
(C)
HexCer Y/X:0
HexCer Y/X:1
Sphingoid base
18:2;O2
18:1;O2
16:1;O2
18:1;O2
16
18
20
22
23
24
24
log2 (FC T/N)
1.5
1.0
0.5
0.0
-0.5
-1.0
-1.5
-log10 (p-value)
2.0
3.0
4.0
5.0
6.0
7.0
8.0
Number of carbons in N-acyl chain

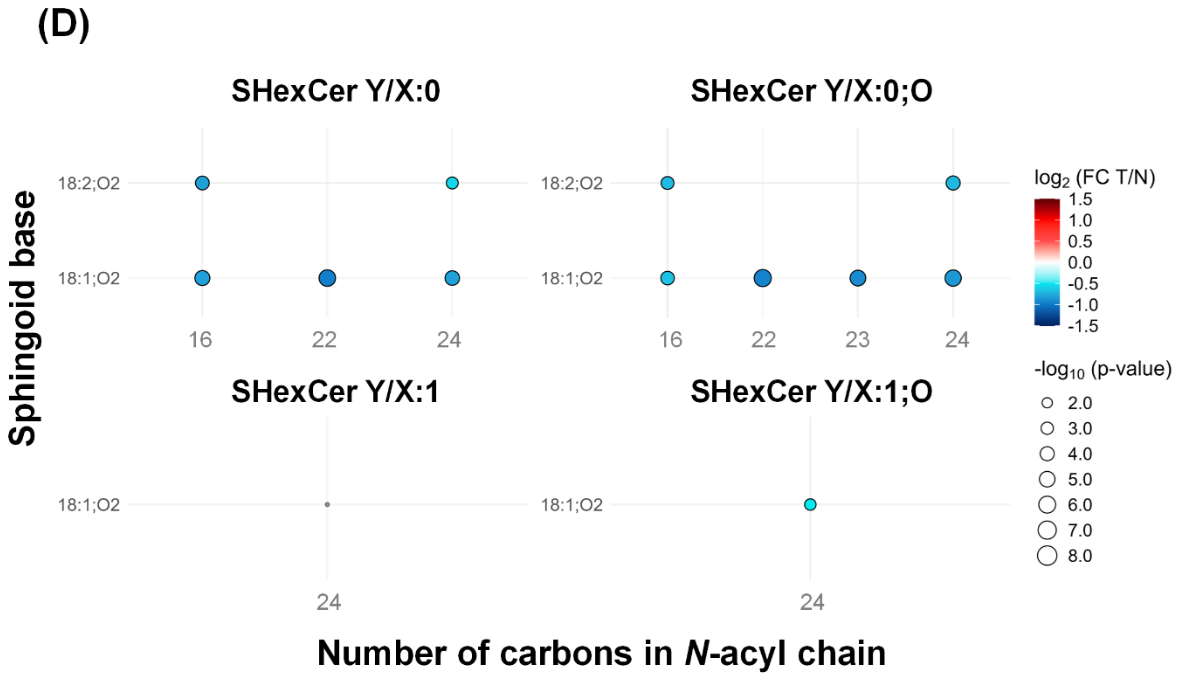
(D)
SHexCer Y/X:0
SHexCer Y/X:0;O
Sphingoid base
18:2;O2
18:1;O2
18:2;O2
18:1;O2
16
22
24
16
22
23
24
SHexCer Y/X:1
SHexCer Y/X:1;O
18:1;O2
18:1;O2
24
24
log2 (FC T/N)
1.5
1.0
0.5
0.0
-0.5
-1.0
-1.5
-log10 (p-value)
2.0
3.0
4.0
5.0
6.0
7.0
8.0
Number of carbons in N-acyl chain

**Figure S10:** Comparison of statistical significance between *sn-1* and *sn-2* isomers of lysophospholipids. Significances were determined using the Mann–Whitney U test with the following thresholds: ns ($p > 0.05$), * ($p \leq 0.05$), ** ($p \leq 0.01$), *** ($p \leq 0.001$), and **** ($p \leq 0.0001$). N... healthy controls (blue); T... PDAC patients (red).

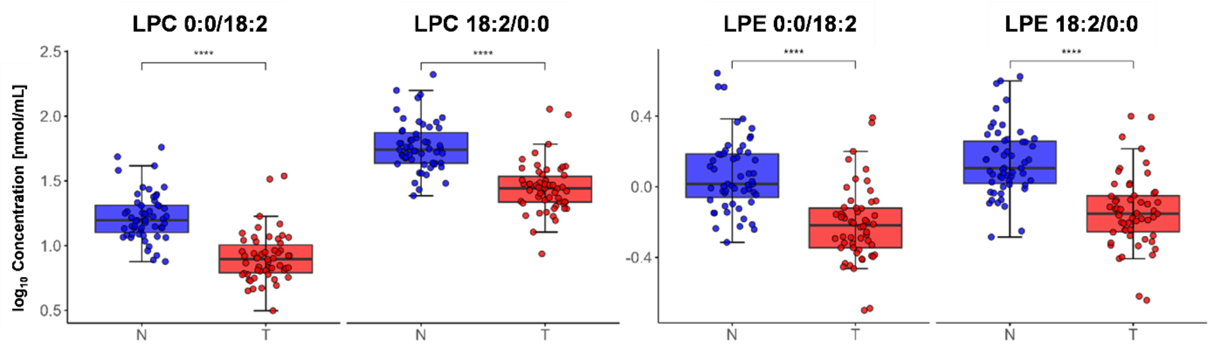

**Figure S11:** Box plots of the most dysregulated phospholipids. Significances were determined using the Mann–Whitney U test with the following thresholds: ns ($p > 0.05$), * ($p \leq 0.05$), ** ($p \leq 0.01$), *** ($p \leq 0.001$), and **** ($p \leq 0.0001$). N... healthy controls (blue); T... PDAC patients (red).

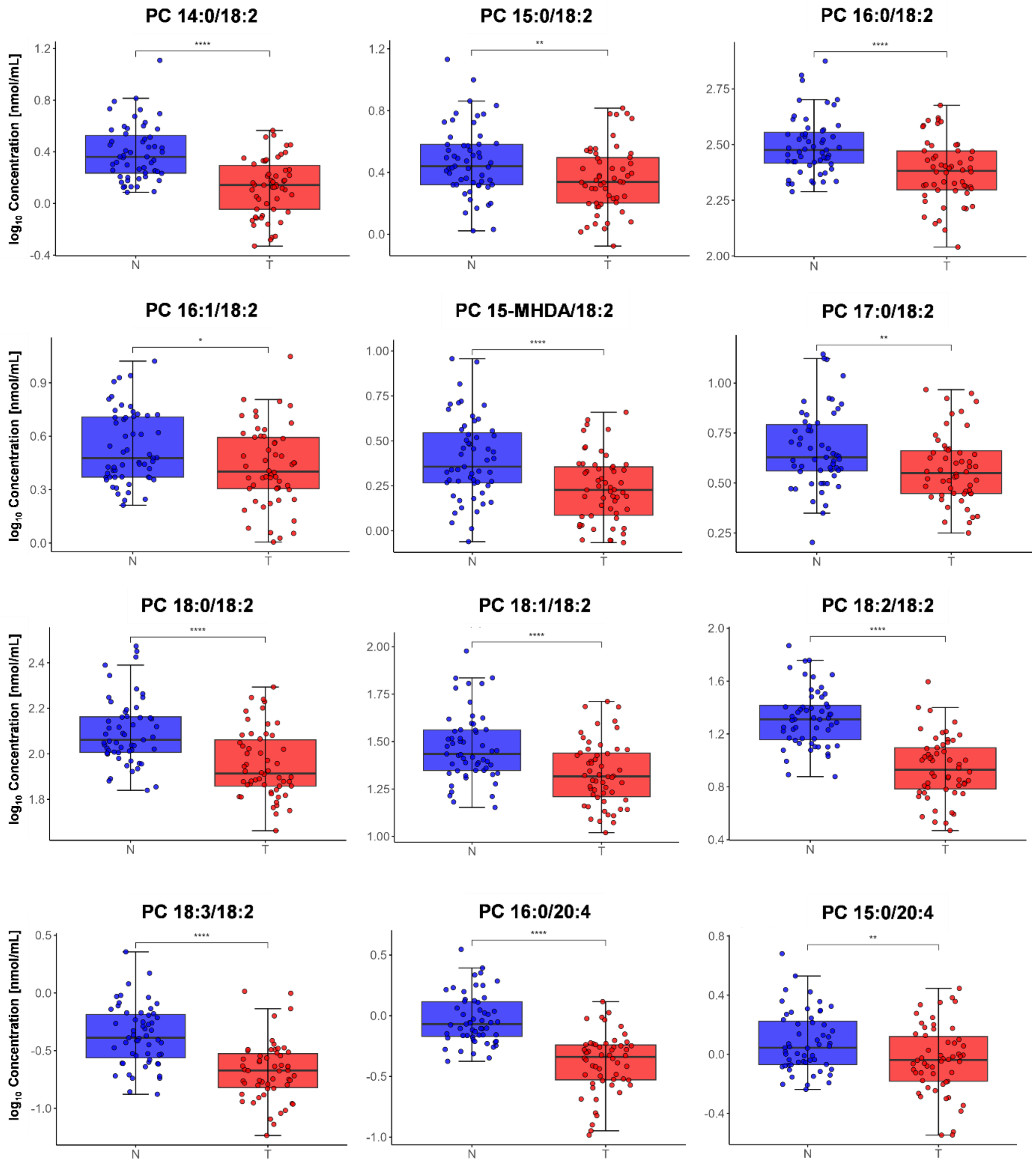

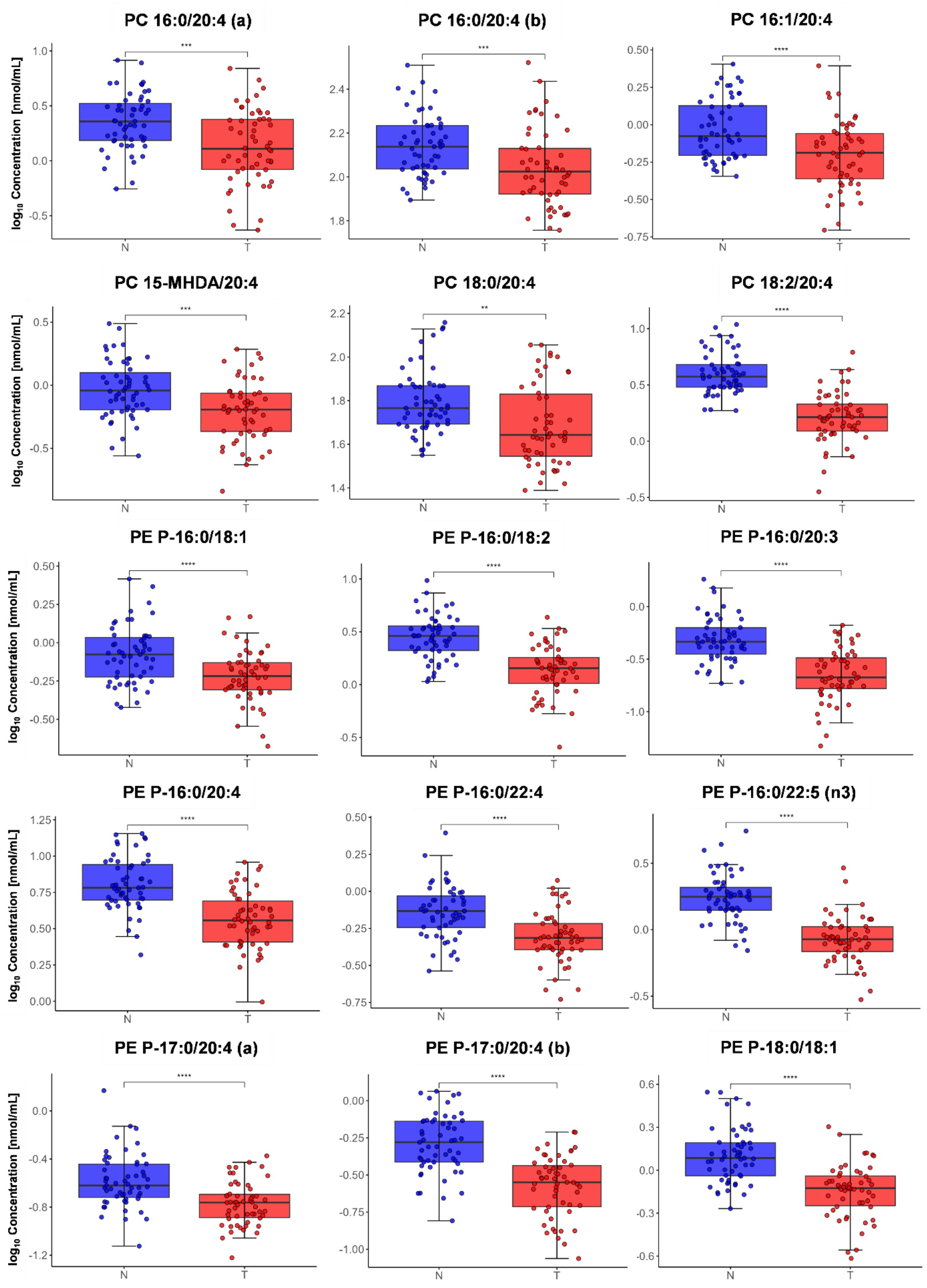
PC 16:0/20:4 (a)
PC 16:0/20:4 (b)
PC 16:1/20:4
PC 15-MHDA/20:4
PC 18:0/20:4
PC 18:2/20:4
PE P-16:0/18:1
PE P-16:0/18:2
PE P-16:0/20:3
PE P-16:0/20:4
PE P-16:0/22:4
PE P-16:0/22:5 (n3)
PE P-17:0/20:4 (a)
PE P-17:0/20:4 (b)
PE P-18:0/18:1
log10 Concentration [nmol/mL]
N
T
***
**
****

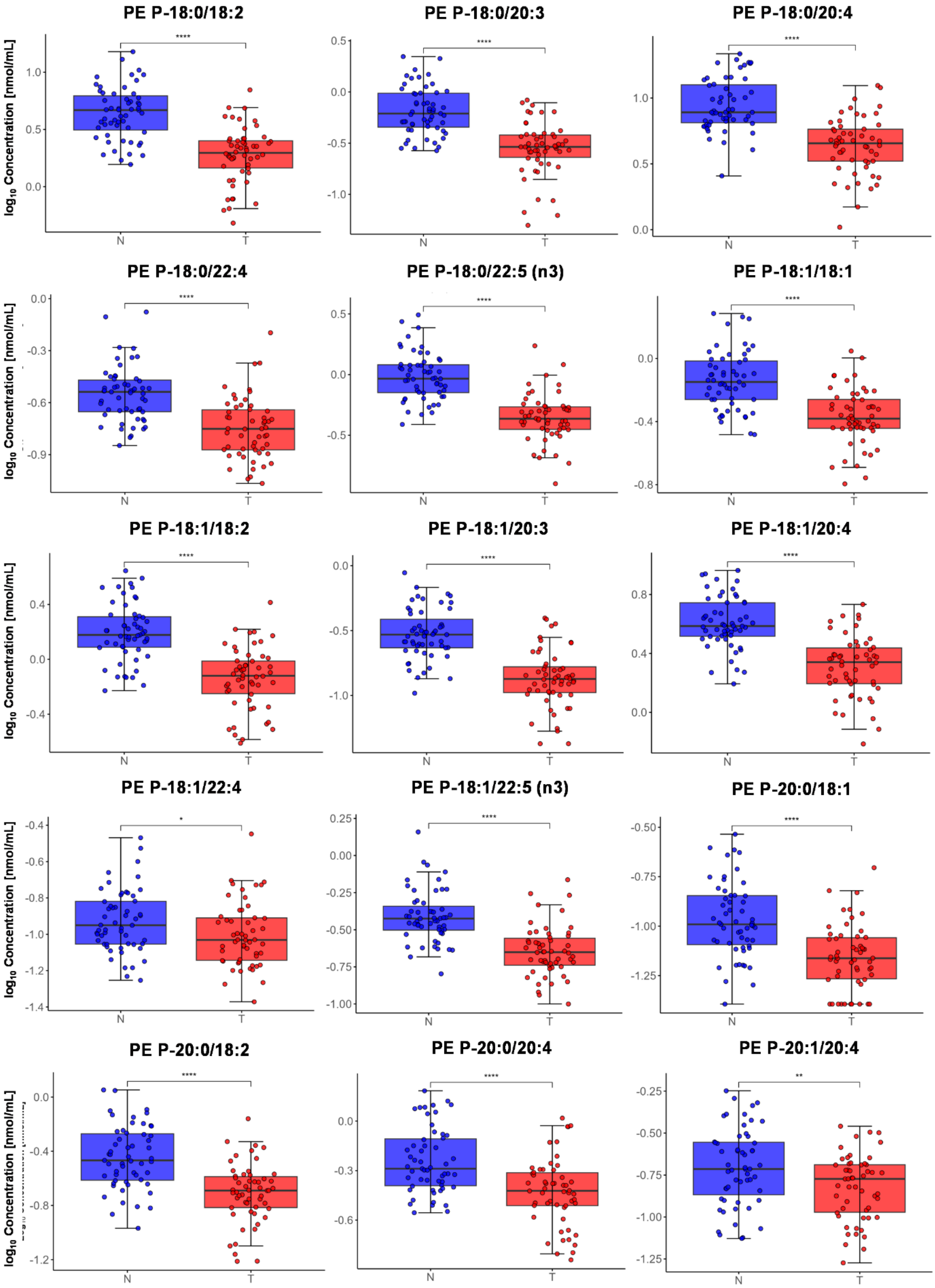

PE P-18:0/18:2
****
PE P-18:0/20:3
****
PE P-18:0/20:4
****
PE P-18:0/22:4
****
PE P-18:0/22:5 (n3)
****
PE P-18:1/18:1
****
PE P-18:1/18:2
****
PE P-18:1/20:3
****
PE P-18:1/20:4
****
PE P-18:1/22:4
*
PE P-18:1/22:5 (n3)
****
PE P-20:0/18:1
****
PE P-20:0/18:2
****
PE P-20:0/20:4
****
PE P-20:1/20:4
**
log10 Concentration [nmol/mL]
N
T

**Figure S12:** Total ion chromatotogram of FAMEs in human serum, measured by GC/MS.

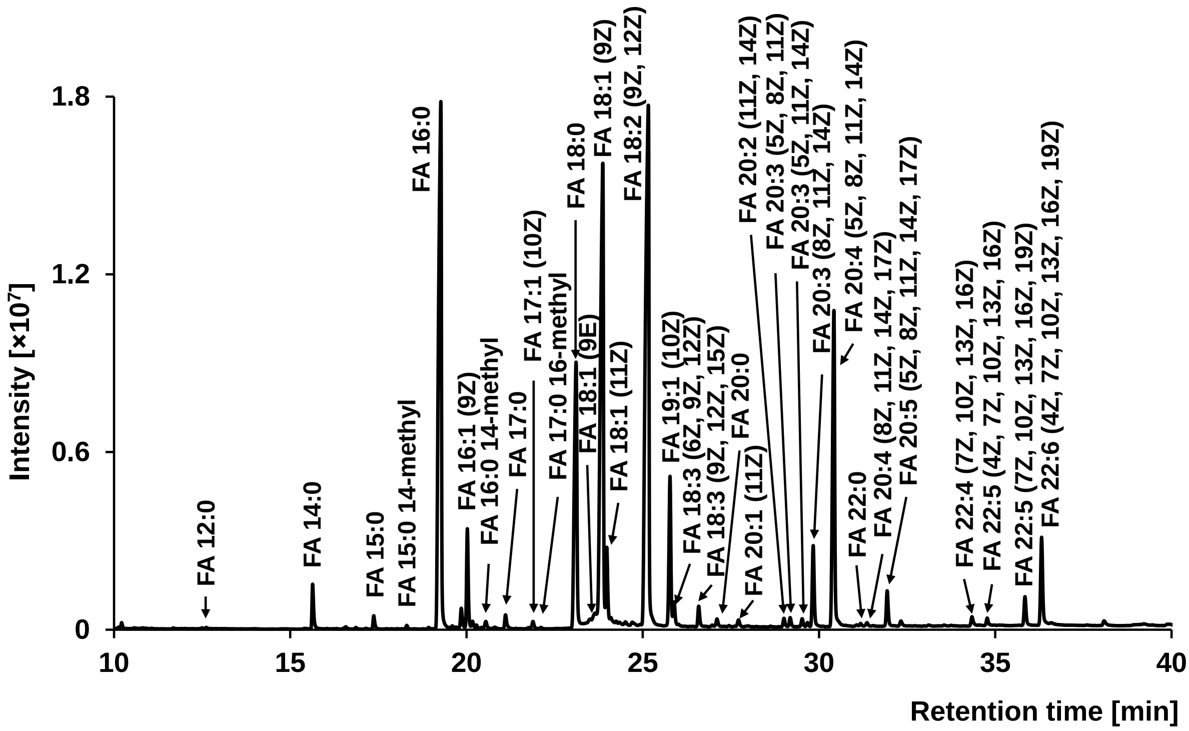

**Figure S13:** S-plot generated from OPLS-DA showing the most upregulated (red) and the most downregulated (blue) FAMEs in PDAC.

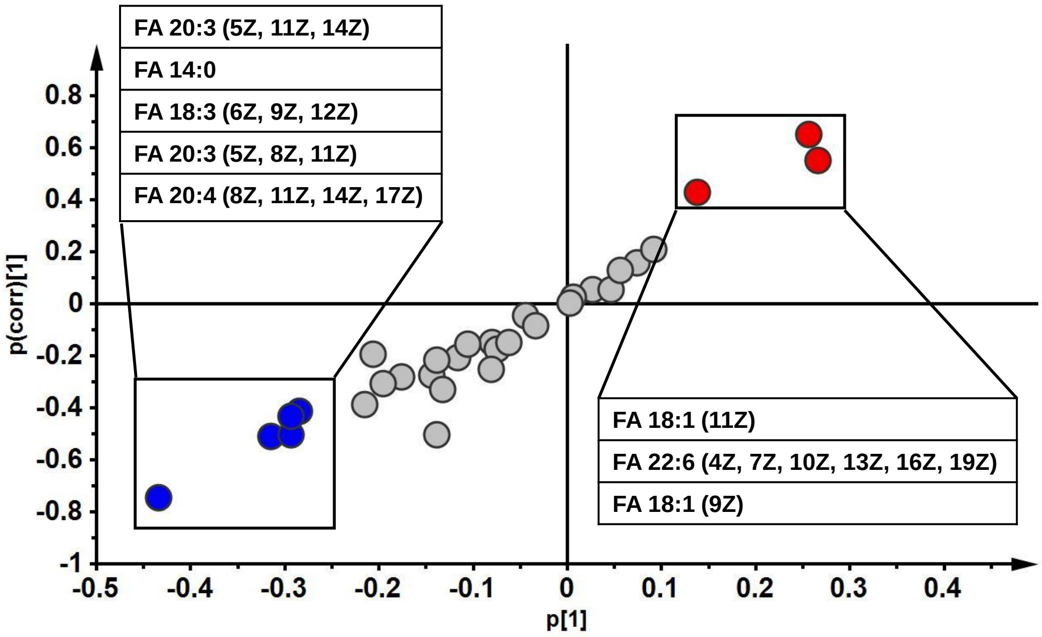

**Figure S14:** Box plots of the most dysregulated FAMEs. Significances were determined using the Mann–Whitney U test with the following thresholds: ns ($p > 0.05$), * ($p \leq 0.05$), ** ($p \leq 0.01$), *** ($p \leq 0.001$), and **** ($p \leq 0.0001$). N... healthy controls (blue); T... PDAC patients (red).

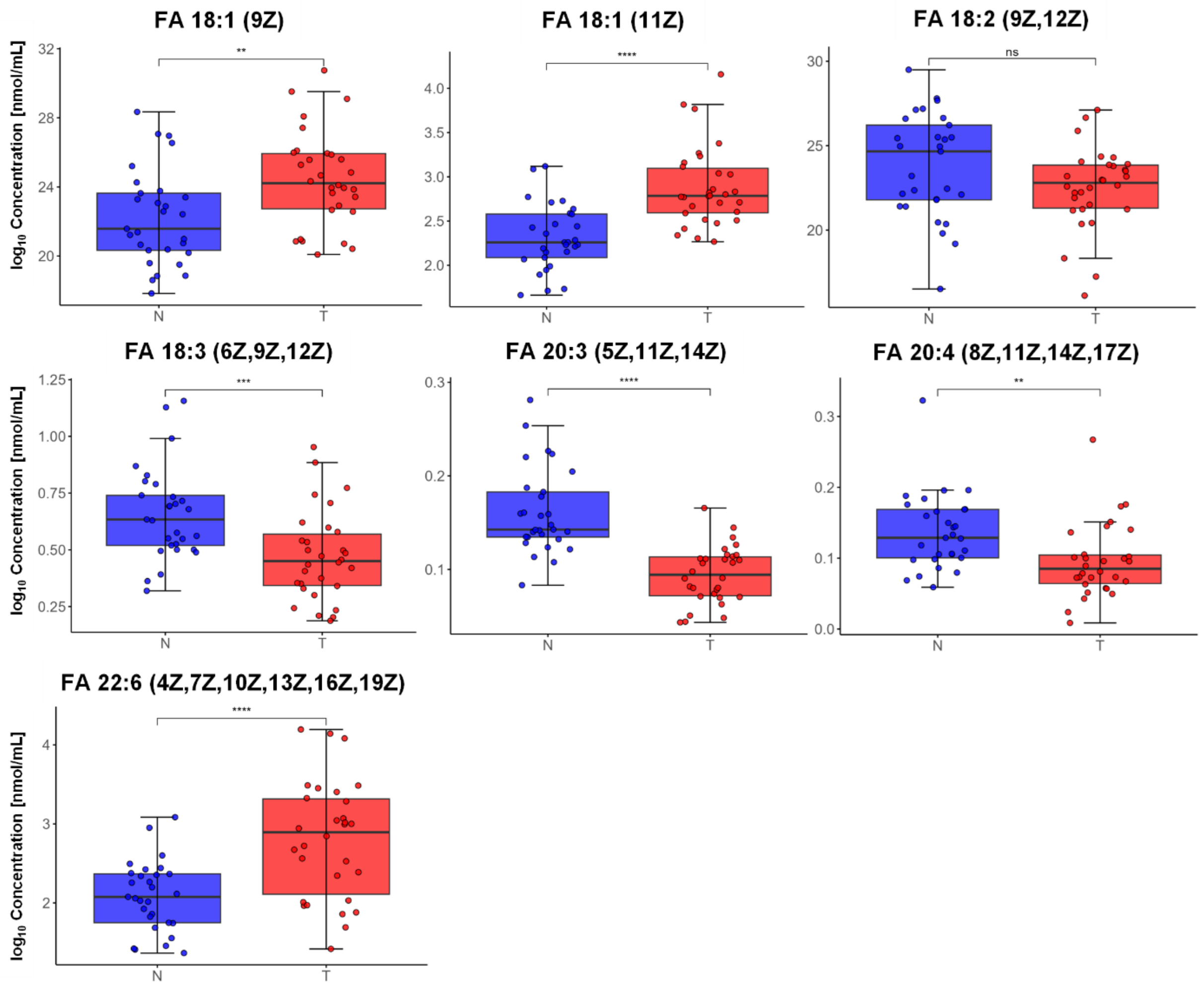
